# Streamlined point-of-care ultrasound examination for pulmonary tuberculosis

**DOI:** 10.64898/2026.11.09.26367649

**Authors:** Lydia Nakiyingi, Rohan Dhamdhere, Wycliff Nate, Keith Guevarra, Eva Nabawanuka, Elena Jovanovska Janeva, Raphael Shankman, Eria Lwanga, Elizabeth Nakabugo, Emmanuel Nasinghe, Sarah Nakayenga, Guadalupe Astorga, Thejaswini Anapakula Sridhar, Hyndavi Krishna Javvaji, Emily Clark, Deborah Handler, Padmini Salgame, David Alland, Moses Joloba, Jerrold Ellner, Anant Madabhushi, Yingda L. Xie

## Abstract

**Background:** Point-of-care ultrasound (POCUS) is a low-cost, radiation-free triage tool but lacks standardized acquisition protocols for tuberculosis (TB). We developed and evaluated a streamlined symmetric POCUS protocol for pulmonary TB in a high-burden setting.

**Methods:** Ugandan participants aged ≥13 years with confirmed pulmonary TB, household TB exposure, or a non-TB respiratory condition underwent an 18-position POCUS examination (12 lung zones and 6 pre-established extrapulmonary positions). Random Forest classifiers using prespecified pathologic features were developed in a fixed stratified 75/25 training/holdout split. All symmetric 2-, 4-, and 6-zone combinations were evaluated in training data. The best protocol of each size was locked and tested in the holdout set.

**Results:** Among 318 analyzed participants, 118 (37.1%) had pulmonary TB and 200 were TB-negative, including 102 with other respiratory conditions (ORC). The model-selected 6-zone protocol comprised bilateral anterior-superior, lateral-superior, and posterior-superior lung zones. For TB versus all TB-negative participants, the AUCs for the 6-zone, 12-zone, and 18-position protocols were 0.82 (95% CI, 0.73-0.92), 0.81 (0.70–0.92), and 0.81 (0.71–0.91), respectively (all P > 0.76). For TB versus ORC, AUCs were 0.81 (0.70–0.91), 0.85 (0.76–0.93), and 0.85 (0.76–0.94), respectively (all P > 0.45). The 2-zone protocol had a lower AUC than the 6-zone protocol for TB versus all TB-negative participants (*P* = 0.004). Shred sign, consolidation, and B-lines with irregular pleura were the strongest TB-associated findings.

**Conclusions:** A fixed 6-zone lung POCUS examination showed similar discrimination for TB to more comprehensive 12-lung-zone and 18-position protocols. This supports a simpler, scalable approach to TB triage.

## INTRODUCTION

Despite progress in tuberculosis (TB) control, more than one in five incident TB cases are not diagnosed or reported each year, reflecting the persistent gap between estimated incidence and case notifications, particularly in high-burden, resource-limited settings.^1–3^ Limited access to chest radiography in Uganda supports the need for portable TB screening tools.^4^ High-throughput screening is constrained by the operational demands and costs of molecular assays and chest radiography, underscoring the need for affordable, field-deployable triage tools that can be deployed at scale.^3,5–7^

Point-of-care ultrasound (POCUS) offers a portable, radiation-free, and relatively low-cost alternative that can be performed by non-specialist healthcare workers in community settings.^8–10^ However, its broader adoption is limited by operator dependence, heterogeneous image acquisition, and lack of standardized acquisition protocols, which may undermine reproducibility and scalability.^8,10^ The Focused Assessment with Sonography for HIV-associated tuberculosis (FASH) protocol focuses on extrapulmonary manifestations of HIV-associated TB,^9,11^ while the BLUE protocol uses standardized lung ultrasound points for evaluating acute respiratory failure^12^; neither was designed to optimize lung scanning for pulmonary TB detection

For pulmonary TB, lung ultrasound (LUS) can detect peripheral abnormalities particularly subpleural nodules and small consolidations, with reported sensitivities generally ranging from approximately 70% to >90%.^10,13–16^ However, in these studies, the specificity of small subpleural consolidations was generally low (16%–39%), reflecting substantial overlap with other respiratory conditions.^14–16^ In Papua New Guinea, for example, a comprehensive LUS examination achieved 92.6% sensitivity and 46.8% specificity for pulmonary TB, whereas a structured score incorporating multiple sonographic features improved specificity to 84.2% at a modestly reduced sensitivity of 82.8%.^15^ These observations support multivariable, pattern-based approaches, including machine learning models that can identify the combination of pathologic features—and potentially the subset of scanning sites—most informative for TB detection. ^14, 15, 17^

We aimed to derive and internally evaluate a streamlined lung POCUS protocol for pulmonary TB that could reduce scanning burden while preserving discrimination for identifying individuals who should be prioritized for confirmatory TB testing.

## MATERIAL AND METHODS

### Study Population and Reference Standard

In this prospective Uganda TBRU/Halting TB sub-study, participants aged 13 years or older were recruited as outpatients through KCC clinics from July 2023 to March 2025. Groups included microbiologically-confirmed pulmonary TB index cases (ICs), household contacts (HHCs), and TB-negative symptomatic participants with another respiratory condition (ORC). IC eligibility and final analytic pulmonary TB status was based on sputum Xpert MTB/RIF Ultra (Ultra) and mycobacterial culture results. Ultra trace results required culture confirmation (e-Table 1). Participants provided consent or, for minors, assent with parental/guardian permission. Exclusions for severe illness, likely loss to follow-up, and TB treatment beyond protocol limits are detailed in the Supplementary Methods.

### POCUS Acquisition and Image Interpretation

All participants underwent a standardized 18-position POCUS examination comprising 12 lung zones and six FASH positions (Figure 1). Examinations used a Philips Lumify phased-array transducer connected to an Android tablet. Six-second cine loops were recorded at each position.

**Figure 1.**
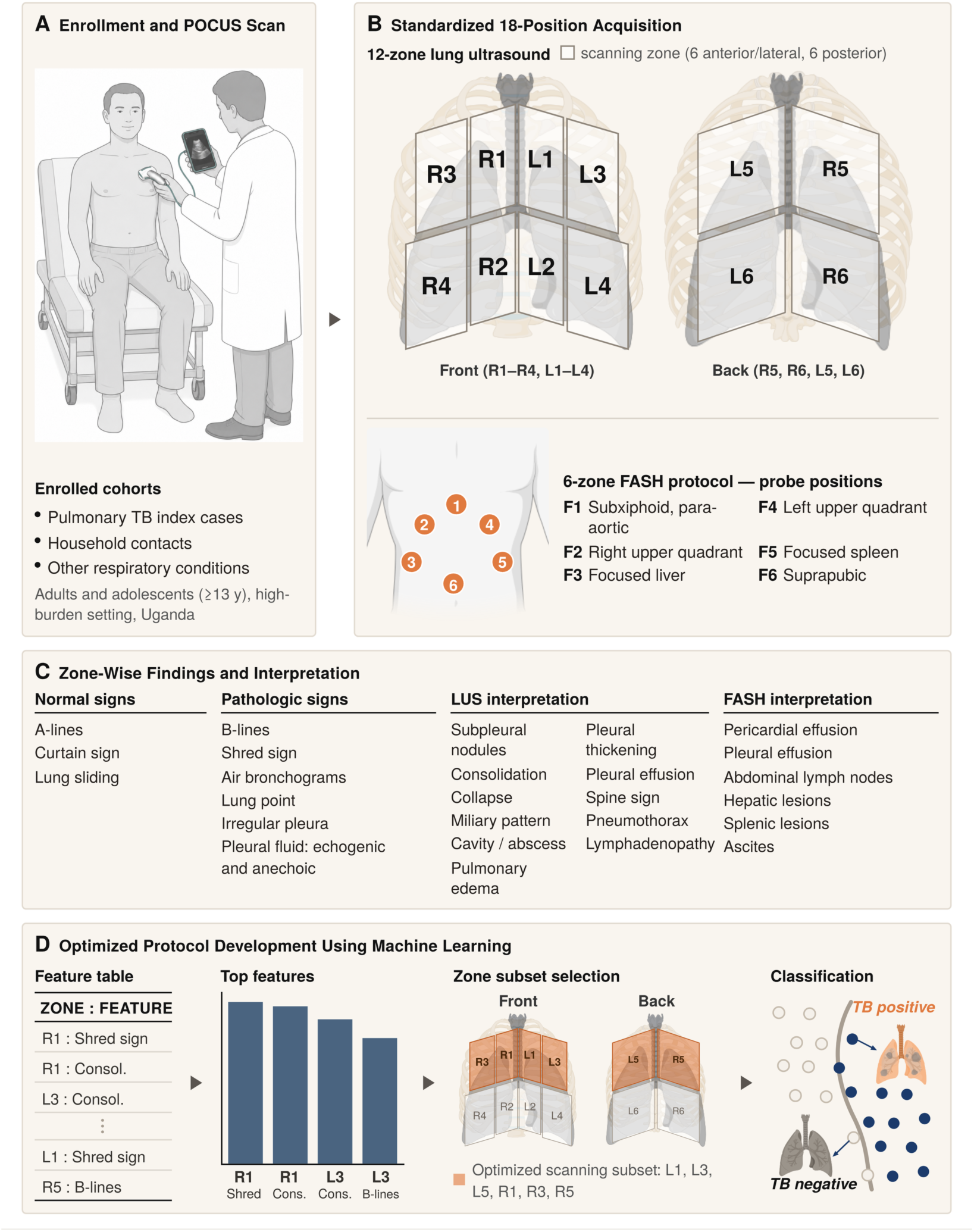
Study workflow and derivation of streamlined POCUS protocols. (A) Enrollment and POCUS examination of pulmonary TB index cases, household contacts, and participants with other respiratory conditions. (B) Standardized 18-position acquisition comprising 12 lung zones and six FASH positions. (C) Findings from lung zones and FASH positions recorded for analysis. (D) Machine-learning development of reduced protocols, yielding the fixed 6-zone set L1, L3, L5, R1, R3, and R5.

Professionally qualified sonographers were trained by expert clinicians - a pulmonologist and radiologist with expertise in ultrasonography. Training consisted of an initial 1-hour didactic session and 2-hour virtual demonstration on a live model, followed by successive rounds of instruction and feedback during the study. For each examination, the sonographer prospectively recorded an interpretation of pathologic findings. The expert clinicians then independently reviewed the stored cine loops, and any disagreements were resolved by feedback and consensus with the sonographer. Sonographers and reviewers were fully blinded to participants’ TB status and enrollment status. Detailed procedures are provided in the Supplementary Methods.

### Chest Radiography

Protocol-specified chest radiographs (CXRs) were obtained for ICs and HHCs, but not ORC participants. A separate pulmonologist retrospectively interpreted frontal and lateral images using a predefined case report form. The pulmonologist was blinded to individual participant TB status and POCUS findings but aware of the overall number TB cases in the cohort, thus blinding of CXR reads was partial.

### Machine Learning Derivation and Evaluation of Streamlined POCUS Protocols

We developed models to identify reduced POCUS protocols for pulmonary TB detection. Models classified TB versus all TB-negative participants (N = 318; training, n = 238; holdout, n = 80, including 30 with TB) and TB versus ORC (N = 220; training, n = 165; holdout, n = 55, including 29 with TB). TB versus participants with no respiratory condition (NRC) was prespecified as a supplemental analysis (N = 216; training, n = 162; holdout, n = 54, including 29 with TB). Random forest (RF) classifiers were trained using complete-case REDCap data without imputation.

Candidate predictors were subpleural nodules, consolidation, B-lines with irregular or smooth pleura, echogenic or anechoic pleural fluid, and shred sign (e-Table 2). The two B-line predictors combined B-lines with the corresponding pleural appearance. Signs of normal aeration were excluded. Cavity or abscess was excluded because no positive findings were recorded. Feature definitions, exclusions, and FASH predictors are detailed in the Supplementary Methods.

Within each comparison, participants were assigned to a fixed 75:25 training/holdout split stratified by TB status; the same split was used for every protocol. All symmetric 2-, 4-, and 6-zone lung combinations were compared with complete 12-zone lung and 18-position lung-plus-FASH protocols.

Protocols were selected using stratified fivefold cross-validation repeated 10 times within the training set. Preprocessing, feature selection, and the synthetic minority oversampling technique (SMOTE) were performed only in each fold’s training subset. For each zone count, we selected the protocol with the highest mean cross-validation area under the receiver operating characteristic curve (AUC) for TB versus all TB-negative participants. Selected zones were used unchanged for the other comparisons. Holdout sets were used for evaluation. Modeling details appear in the Supplementary Methods.

AUC confidence intervals and paired comparisons used DeLong’s method. Sensitivity was estimated at 60% and 70% specificity, with 95% CIs from 1,000 bootstrap samples. Other classification metrics used the threshold maximizing Youden’s index in the holdout set. Feature-enrichment tests and multiple-testing adjustments are detailed in the Supplementary Methods.

Exploratory analyses by HIV and sputum-smear status used holdout predictions without model refitting or additional feature selection. Some subgroups included only five or six TB-positive participants, limiting precision (Supplementary Methods).

### Nomogram Modeling and Statistical analyses

Logistic-regression nomograms used RF-selected 6-zone features, training-only imputation, observed training prevalence without SMOTE, and L2 penalization. Complete-separation predictors were excluded. Fitting criteria are in the Supplementary Methods. Evaluation included AUC, sensitivity at 70% and 60% specificity corresponding to World Health Organization targets,^18,19^ Brier score, calibration intercept, and calibration slope.

Clinical characteristics were compared using Wilcoxon rank-sum, chi-square, or Fisher exact tests in Stata 18.0. Tests were two-sided, with *P* < 0.05 considered significant. Radiographic comparisons were restricted to participants with protocol chest radiography. Supplementary Methods provide statistical procedures, POCUS-positive definitions, and anatomical-correspondence rules.

## RESULTS

### Participant Characteristics

A total of 407 participants were screened, of whom 84 were excluded because of ineligibility (n = 33, 31 aged < 13 y and 2 with prior anti-tuberculosis treatment per protocol) or declined participation (n = 51). Of 323 enrolled participants, 5 HHCs were excluded because TB status was indeterminate or POCUS data were incomplete. The final analysis population were 318 participants: 114 ICs, 102 HHCs, and 102 symptomatic clinic participants with ORC (e-Figure 1).

Pulmonary TB was diagnosed in 118 participants (37.1%), comprising 114 ICs and 4 HHCs; 90 of 118 (76.3%) were smear positive. The 200 TB-negative participants comprised of 98 HHCs with no respiratory condition (NRC) and 102 participants with ORC.

Baseline characteristics are shown in Table 1. Participants with pulmonary TB (TB-positive) were younger (median age 31.5 vs 34.5 years in the ORC group, P = .026). BMI was significantly lower in TB-positive individuals (median 19.4) than in the NRC (21.6) and ORC (23.2) groups (both P < .001). HIV prevalence was lowest in the NRC group (15.3%) and similar between TB-positive (24.6%) and ORC (26.5%) participants (P = .09 and P = .75, respectively). Nearly all TB-positive participants (97.5%) reported multiple TB symptoms, compared with 17.3% in the TB-negative NRC group (P < .001) and 88.2% in the ORC group (P = .024).

**Table 1.**
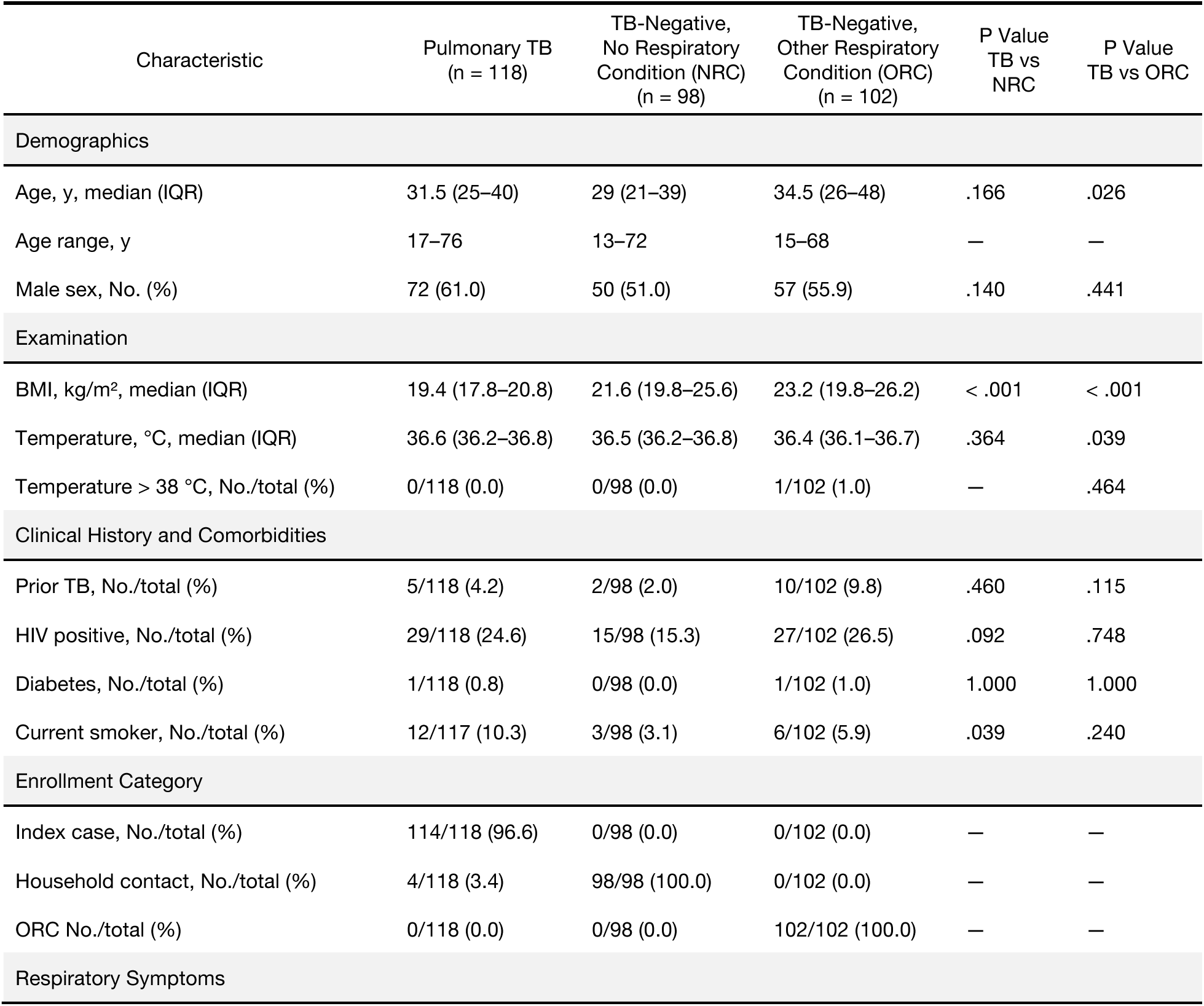
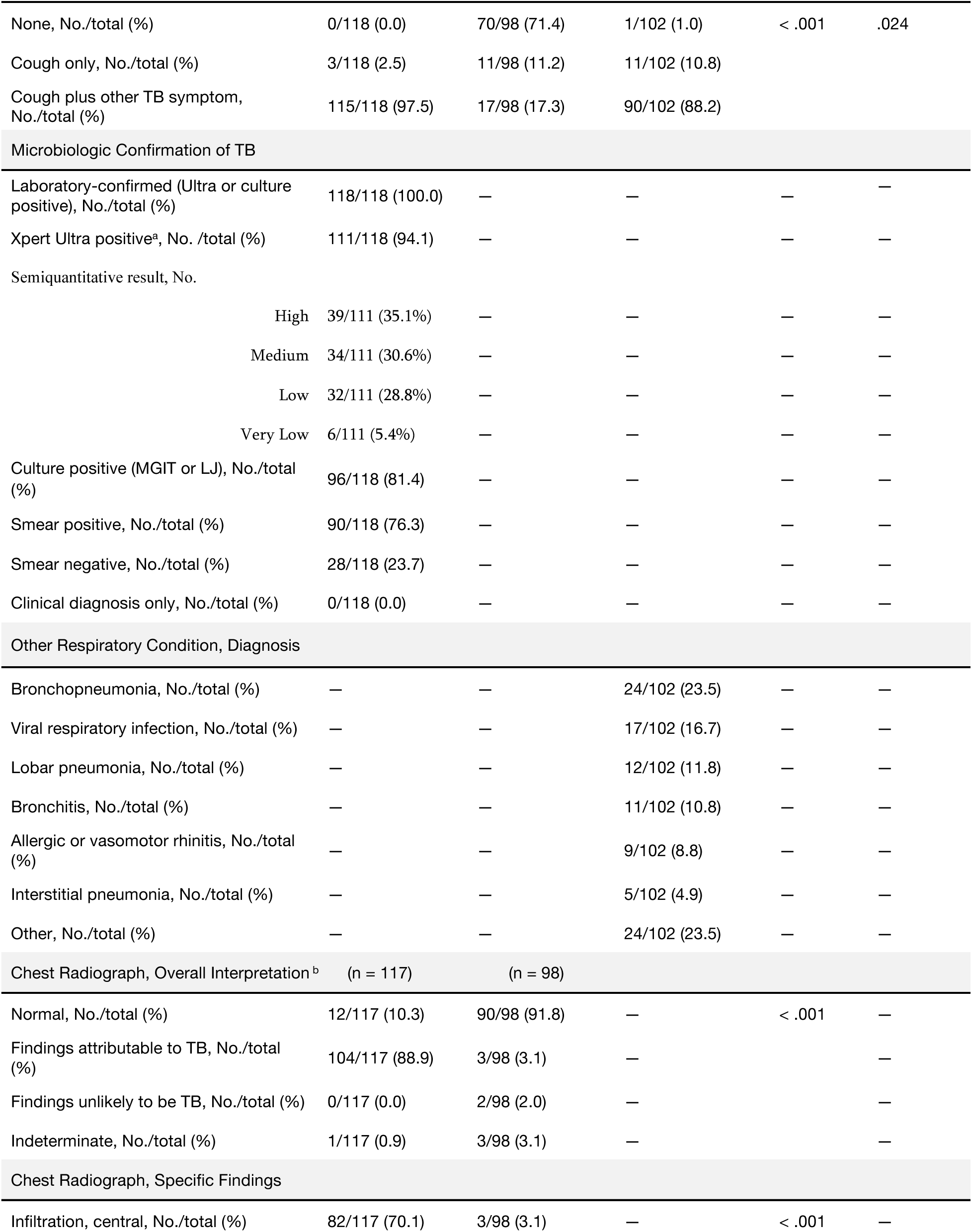
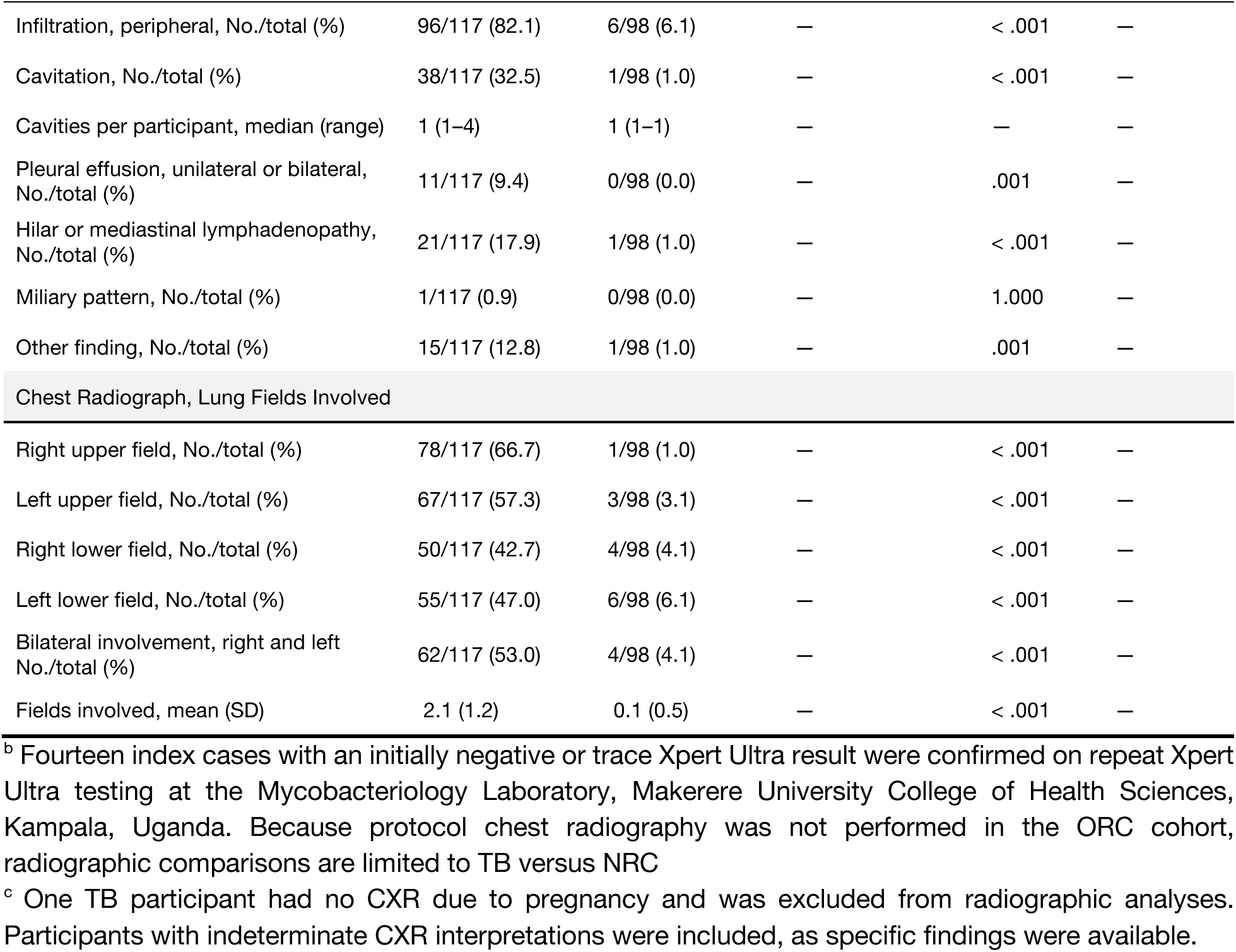
Baseline characteristics of the analyzed cohort by TB status. Data are presented as median (IQR), median (range), range, or No./total (%), as indicated. Pairwise *P* values compare TB with NRC and ORC using Wilcoxon rank-sum, Pearson χ², or Fisher exact tests, as appropriate; tests were two-sided without multiplicity adjustment. TB status was microbiologically determined as detailed in e-Table 1. Microbiologic categories are not mutually exclusive; participants could be positive by more than one method, so component rows do not sum to the total. ORC diagnoses were assigned clinically; viral respiratory infection includes influenza (n = 11) and common cold (n = 6), and bronchitis includes acute (n = 7), chronic (n = 3), and unspecified bronchitis (n = 1). An em dash indicates a comparison that was not applicable or estimable. **Abbreviations:** BMI = body mass index; IQR = interquartile range; LJ = Löwenstein–Jensen; MGIT = Mycobacteria Growth Indicator Tube; NRC = no respiratory condition; ORC = other respiratory condition; TB = tuberculosis; Ultra = Xpert MTB/RIF Ultra.

| Characteristic | Pulmonary TB<br>(n = 118) | TB-Negative,<br>No Respiratory<br>Condition (NRC)<br>(n = 98) | TB-Negative,<br>Other Respiratory<br>Condition (ORC)<br>(n = 102) | P Value<br>TB vs<br>NRC | P Value<br>TB vs ORC |
|---|---|---|---|---|---|
| Demographics |  |  |  |  |  |
| Age, y, median (IQR) | 31.5 (25–40) | 29 (21–39) | 34.5 (26–48) | .166 | .026 |
| Age range, y | 17–76 | 13–72 | 15–68 | — | — |
| Male sex, No. (%) | 72 (61.0) | 50 (51.0) | 57 (55.9) | .140 | .441 |
| Examination |  |  |  |  |  |
| BMI, kg/m<sup>2</sup>, median (IQR) | 19.4 (17.8–20.8) | 21.6 (19.8–25.6) | 23.2 (19.8–26.2) | < .001 | < .001 |
| Temperature, °C, median (IQR) | 36.6 (36.2–36.8) | 36.5 (36.2–36.8) | 36.4 (36.1–36.7) | .364 | .039 |
| Temperature > 38 °C, No./total (%) | 0/118 (0.0) | 0/98 (0.0) | 1/102 (1.0) | — | .464 |
| Clinical History and Comorbidities |  |  |  |  |  |
| Prior TB, No./total (%) | 5/118 (4.2) | 2/98 (2.0) | 10/102 (9.8) | .460 | .115 |
| HIV positive, No./total (%) | 29/118 (24.6) | 15/98 (15.3) | 27/102 (26.5) | .092 | .748 |
| Diabetes, No./total (%) | 1/118 (0.8) | 0/98 (0.0) | 1/102 (1.0) | 1.000 | 1.000 |
| Current smoker, No./total (%) | 12/117 (10.3) | 3/98 (3.1) | 6/102 (5.9) | .039 | .240 |
| Enrollment Category |  |  |  |  |  |
| Index case, No./total (%) | 114/118 (96.6) | 0/98 (0.0) | 0/102 (0.0) | — | — |
| Household contact, No./total (%) | 4/118 (3.4) | 98/98 (100.0) | 0/102 (0.0) | — | — |
| ORC No./total (%) | 0/118 (0.0) | 0/98 (0.0) | 102/102 (100.0) | — | — |
| Respiratory Symptoms |  |  |  |  |  |
| None, No./total (%) | 0/118 (0.0) | 70/98 (71.4) | 1/102 (1.0) | < .001 | .024 |
| Cough only, No./total (%) | 3/118 (2.5) | 11/98 (11.2) | 11/102 (10.8) |  |  |
| Cough plus other TB symptom, No./total (%) | 115/118 (97.5) | 17/98 (17.3) | 90/102 (88.2) |  |  |
| Microbiologic Confirmation of TB |  |  |  |  |  |
| Laboratory-confirmed (Ultra or culture positive), No./total (%) | 118/118 (100.0) | — | — | — | — |
| Xpert Ultra positive<sup>a</sup>, No. /total (%) | 111/118 (94.1) | — | — | — | — |
| Semiquantitative result, No. |  |  |  |  |  |
| High | 39/111 (35.1%) | — | — | — | — |
| Medium | 34/111 (30.6%) | — | — | — | — |
| Low | 32/111 (28.8%) | — | — | — | — |
| Very Low | 6/111 (5.4%) | — | — | — | — |
| Culture positive (MGIT or LJ), No./total (%) | 96/118 (81.4) | — | — | — | — |
| Smear positive, No./total (%) | 90/118 (76.3) | — | — | — | — |
| Smear negative, No./total (%) | 28/118 (23.7) | — | — | — | — |
| Clinical diagnosis only, No./total (%) | 0/118 (0.0) | — | — | — | — |
| Other Respiratory Condition, Diagnosis |  |  |  |  |  |
| Bronchopneumonia, No./total (%) | — | — | 24/102 (23.5) | — | — |
| Viral respiratory infection, No./total (%) | — | — | 17/102 (16.7) | — | — |
| Lobar pneumonia, No./total (%) | — | — | 12/102 (11.8) | — | — |
| Bronchitis, No./total (%) | — | — | 11/102 (10.8) | — | — |
| Allergic or vasomotor rhinitis, No./total (%) | — | — | 9/102 (8.8) | — | — |
| Interstitial pneumonia, No./total (%) | — | — | 5/102 (4.9) | — | — |
| Other, No./total (%) | — | — | 24/102 (23.5) | — | — |
| Chest Radiograph, Overall Interpretation<sup>b</sup> | (n = 117) | (n = 98) |  |  |  |
| Normal, No./total (%) | 12/117 (10.3) | 90/98 (91.8) | — | < .001 | — |
| Findings attributable to TB, No./total (%) | 104/117 (88.9) | 3/98 (3.1) | — |  | — |
| Findings unlikely to be TB, No./total (%) | 0/117 (0.0) | 2/98 (2.0) | — |  | — |
| Indeterminate, No./total (%) | 1/117 (0.9) | 3/98 (3.1) | — |  | — |
| Chest Radiograph, Specific Findings |  |  |  |  |  |
| Infiltration, central, No./total (%) | 82/117 (70.1) | 3/98 (3.1) | — | < .001 | — |
| Infiltration, peripheral, No./total (%) | 96/117 (82.1) | 6/98 (6.1) | — | < .001 | — |
| Cavitation, No./total (%) | 38/117 (32.5) | 1/98 (1.0) | — | < .001 | — |
| Cavities per participant, median (range) | 1 (1–4) | 1 (1–1) | — | — | — |
| Pleural effusion, unilateral or bilateral, No./total (%) | 11/117 (9.4) | 0/98 (0.0) | — | .001 | — |
| Hilar or mediastinal lymphadenopathy, No./total (%) | 21/117 (17.9) | 1/98 (1.0) | — | < .001 | — |
| Miliary pattern, No./total (%) | 1/117 (0.9) | 0/98 (0.0) | — | 1.000 | — |
| Other finding, No./total (%) | 15/117 (12.8) | 1/98 (1.0) | — | .001 | — |
| Chest Radiograph, Lung Fields Involved |  |  |  |  |  |
| Right upper field, No./total (%) | 78/117 (66.7) | 1/98 (1.0) | — | < .001 | — |
| Left upper field, No./total (%) | 67/117 (57.3) | 3/98 (3.1) | — | < .001 | — |
| Right lower field, No./total (%) | 50/117 (42.7) | 4/98 (4.1) | — | < .001 | — |
| Left lower field, No./total (%) | 55/117 (47.0) | 6/98 (6.1) | — | < .001 | — |
| Bilateral involvement, right and left No./total (%) | 62/117 (53.0) | 4/98 (4.1) | — | < .001 | — |
| Fields involved, mean (SD) | 2.1 (1.2) | 0.1 (0.5) | — | < .001 | — |

<sup>b</sup> Fourteen index cases with an initially negative or trace Xpert Ultra result were confirmed on repeat Xpert Ultra testing at the Mycobacteriology Laboratory, Makerere University College of Health Sciences, Kampala, Uganda. Because protocol chest radiography was not performed in the ORC cohort, radiographic comparisons are limited to TB versus NRC
<sup>c</sup> One TB participant had no CXR due to pregnancy and was excluded from radiographic analyses. Participants with indeterminate CXR interpretations were included, as specific findings were available.

Among participants with ORC, the most common diagnoses were bronchopneumonia (23.5%), viral respiratory infection (16.7%), and lobar pneumonia (11.8%).

### Diagnostic Performance of Reduced and Comprehensive POCUS Protocols

The training-data candidate search selected L3/R3 for the 2-zone protocol, L3/L5/R3/R5 for the 4-zone protocol, and L1/L3/L5/R1/R3/R5 for the 6-zone protocol (e-Table 6).

For TB versus all TB-negative participants, the selected 6-zone protocol achieved an AUC of 0.82 (95% CI, 0.73-0.92) and sensitivity of 0.81 (95% CI, 0.68-0.93) at 70% specificity. Its AUC was comparable to that of the 12-zone lung protocol (0.81, 6 vs 12 zones, *P* = 0.7667) and 18-position lung-plus-FASH protocol (0.81, 6 zones vs 18 positions, *P* = 0.7852). The 4-zone AUC was 0.77 (95% CI, 0.66-0.88, 4 vs 6 zones, *P* = 0.2172). The 2-zone AUC was 0.70 (95% CI, 0.60-0.80) and was lower than the 6-zone AUC (*P* = 0.0042) (Figure 2A, e-Tables 4 and 5).

**Figure 2.**
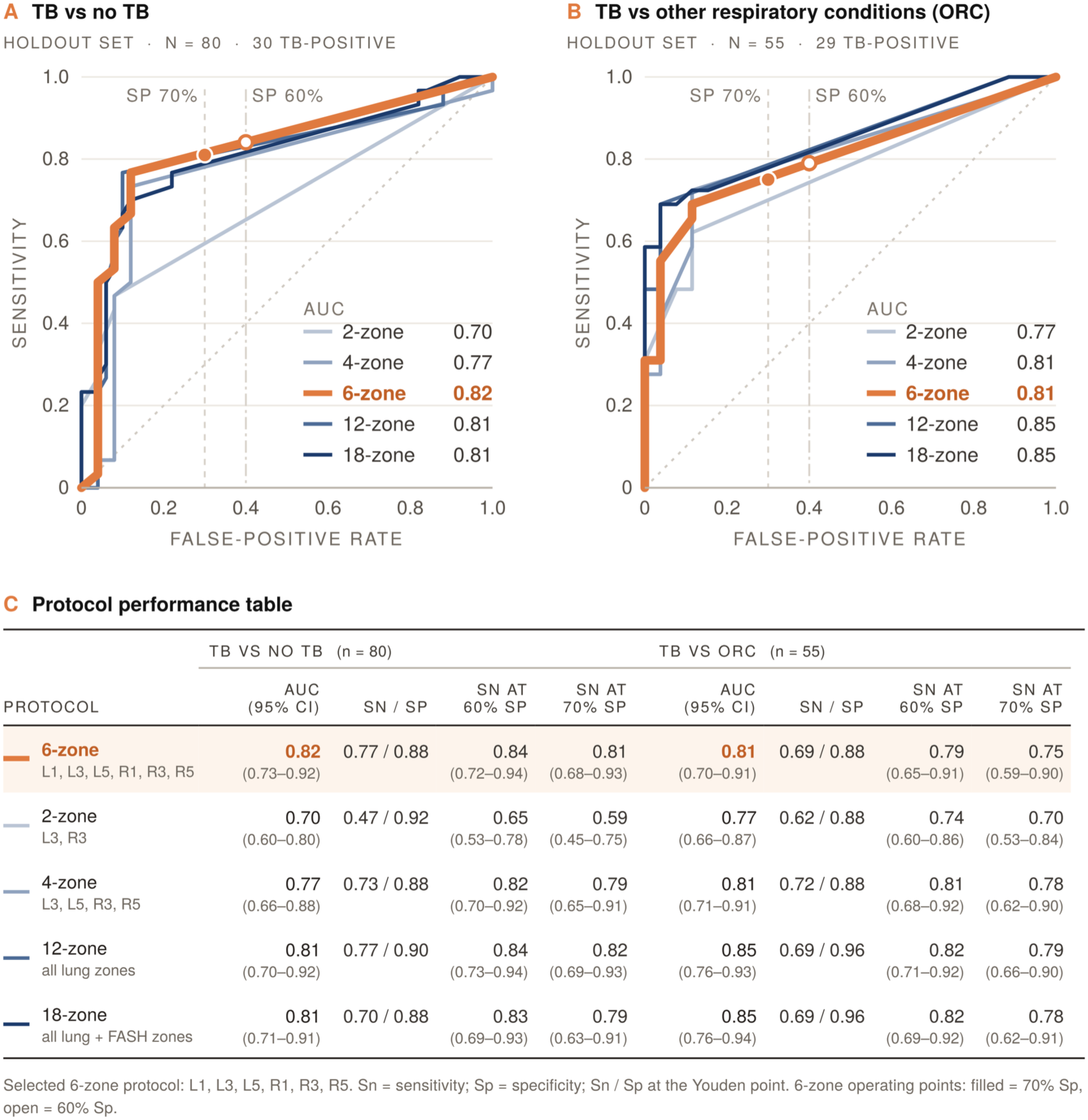
Holdout performance of reduced and comprehensive POCUS protocols: ROC curves compare the 2-, 4-, 6-, and 12-zone lung protocols and the 18-position combined protocol for (A) TB versus all TB-negative participants (N = 80; 30 TB-positive) and (B) TB versus other respiratory conditions (N = 55; 29 TB-positive). (C) Corresponding AUC, sensitivity and specificity at the Youden threshold, and sensitivity at 70% and 60% specificity thresholds. The selected 6-zone protocol comprised L1, L3, L5, R1, R3, and R5.

For TB versus ORC, the 12-zone (AUC 0.85, 95% CI, 0.76-0.93) and 18-position (AUC 0.85, 95% CI, 0.76-0.94) protocols had the highest point estimates, followed by the selected 4-zone and 6-zone protocols (both approximately 0.81) and the 2-zone protocol (0.77). Comparisons of the 6-zone protocol with the 4-zone (*P* = 0.7449), 12-zone (*P* = 0.4556), and 18-position (*P* = 0.4862) protocols were not statistically significant. Sensitivity at 70% specificity was 0.70 for the 2-zone, 0.75 for the 6-zone, and 0.79 for the 12-zone protocol (Figure 2B, e-Tables 4 and 5). Complete TB-versus-NRC performance is reported in e-Table 3.

FASH response variables were retained by the 18-position feature-selection procedure, but adding FASH positions did not improve AUC over the 12-zone lung protocol for TB versus all TB-negative participants (*P* = 0.9439) or TB versus ORC (*P* = 0.9748) (e-Table 5).

### Radiographic Correlates of POCUS Detection

Because CXR was obtained for ICs and HHCs but not the separately enrolled ORC cohort, radiographic comparisons were restricted to TB versus NRC. Central infiltration (70.1% vs 3.1%), peripheral infiltration (82.1% vs 6.1%), cavitation (32.5% vs 1.0%), pleural effusion (9.4% vs 0%), and hilar or mediastinal lymphadenopathy (17.9% vs 1.0%) were more frequent among TB than NRC participants (all P ≤ .001), and 91.8% of NRC participants had normal CXRs (Table 1). POCUS positive TB participants had more extensive radiographic disease than POCUS-negative TB participants, with more frequent central infiltration (79.0% vs 50.0%, P = .002), peripheral infiltration (91.4% vs 61.1%, P < .001), involvement of both upper and lower lung fields (71.6% vs 36.1%, P < .001), and a greater number of involved lung fields (mean 2.4 vs 1.6, P < .001).

Peripheral involvement showed the strongest association with POCUS detection: 74 of 96 participants (77.1%) with peripheral infiltration were POCUS positive, compared with 7 of 21 (33.3%) without peripheral infiltration (P < .001). Cavitation and hilar or mediastinal lymphadenopathy were not directly visualized by POCUS. Among 30 of 38 participants with radiographic cavitation who were POCUS positive, detection was attributed to consolidation (n = 20) and/or subpleural nodules (n = 12), and 29 of 30 (96.7%) also had peripheral infiltration. Notably, 2 of the 117 TB-positive participants were identified by POCUS but had a normal CXR whereas 26 TB participants were POCUS negative but had CXR findings. Overall, 78 of 81 (96.3%) POCUS-positive participants had a TB-suggestive POCUS finding corresponding to a radiographically involved lung field. Full findings are shown in e-Table 12.

### Exploratory Subgroup Analyses

In the exploratory HIV-stratified analyses, estimates were imprecise because each HIV-positive holdout stratum contained only five TB-positive participants. For TB versus all TB-negative participants, the 6-zone AUC was 0.925 (95% CI, 0.766-1.000) among HIV-positive participants and 0.802 (95% CI, 0.695-0.909) among HIV-negative participants. For TB versus ORC, the corresponding 6-zone AUCs were 0.760 (95% CI, 0.467-1.000) and 0.819 (95% CI, 0.704-0.934). 12-zone and 18-position results are provided in e-Table 7. No subgroup superiority or interaction was significant.

In the exploratory analyses of TB stratified by smear-microscopy status, the 6-zone model achieved an AUC of 0.860 (95% CI, 0.685-1.000) and sensitivity of 0.867 (95% CI, 0.586-1.000) at 70% specificity for distinguishing smear-negative TB versus no TB. For smear-positive TB versus no TB, the corresponding AUC was 0.816 (95% CI, 0.713-0.919). Results across the 6-zone, 12-zone, and 18-position models and the TB-versus-ORC endpoint are provided in e-Table 8 and require cautious interpretation because smear-negative strata contained only five or six TB-positive participants.

### Top Zone-Level Pathologic Features

After two-sided Fisher exact testing with Benjamini-Hochberg FDR adjustment across 91 nonconstant processed lung and FASH features, the strongest TB-associated findings were pathologic lung features, particularly shred sign, consolidation, and B-lines with irregular pleura (Table 2). For TB versus all TB-negative participants, the leading features were R1 shred sign, R1 consolidation, and L3 consolidation. For TB versus ORC, the leading features were R3 B-lines with irregular pleura, R1 consolidation, and R1 shred sign. Normal-aeration signs were excluded from this feature ranking. Endpoint-specific RF importance plots are provided in e-Figures 2-4.

**Table 2.** Top 10 zone-level POCUS features in the selected 6-zone modeling workflow. Features are restricted to L1, L3, L5, R1, R3, and R5. For each endpoint, the first five rows correspond to features retained in the moderate nomogram and are listed in the same order as Figure 3; the remaining five are additional RF-retained features with the lowest FDR-adjusted *P* values. FDR-adjusted *P* values were calculated using two-sided Fisher exact tests with Benjamini-Hochberg correction and represent univariate associations, not RF importance or multivariable effects. ***Abbreviations:*** FDR = false discovery rate; ORC = other respiratory conditions; POCUS = point-of-care ultrasound; RF = random forest; TB = tuberculosis.

| **Order** | **TB vs No TB feature** | **FDR-adjusted *P*** | **TB vs ORC feature** | **FDR-adjusted *P*** |
|---|---|---|---|---|
| 1 | L3 — Consolidation | 7.40 × 10<sup>-7</sup> | L3 — B-lines with smooth pleura | 1.44 × 10<sup>-2</sup> |
| 2 | R3 — B-lines with irregular pleura | 1.47 × 10<sup>-4</sup> | R5 — Consolidation | 1.89 × 10<sup>-3</sup> |
| 3 | L5 — B-lines with irregular pleura | 1.11 × 10<sup>-5</sup> | L5 — Consolidation | 2.18 × 10<sup>-3</sup> |
| 4 | R5 — Consolidation | 5.32 × 10<sup>-6</sup> | L3 — B-lines with irregular pleura | 1.99 × 10<sup>-3</sup> |
| 5 | L5 — Consolidation | 7.67 × 10<sup>-6</sup> | L3 — Consolidation | 1.89 × 10<sup>-3</sup> |
| 6 | R1 — Shred sign | 1.27 × 10<sup>-7</sup> | R3 — B-lines with irregular pleura | 1.72 × 10<sup>-4</sup> |
| 7 | R1 — Consolidation | 1.86 × 10<sup>-7</sup> | R1 — Consolidation | 1.72 × 10<sup>-4</sup> |
| 8 | L3 — B-lines with irregular pleura | 4.48 × 10<sup>-6</sup> | R1 — Shred sign | 2.35 × 10<sup>-4</sup> |
| 9 | L1 — B-lines with irregular pleura | 5.32 × 10<sup>-6</sup> | L3 — Shred sign | 1.99 × 10<sup>-3</sup> |
| 10 | L3 — Shred sign | 1.23 × 10<sup>-5</sup> | L1 — Consolidation | 1.99 × 10<sup>-3</sup> |

### Clinically-interpretable Nomograms

Both moderate nomograms used RF-selected predictors from the fixed 6-zone protocol. For TB versus all TB-negative participants, the retained features were L3 consolidation, L5 consolidation, L5 B-lines with irregular pleura, R3 B-lines with irregular pleura, and R5 consolidation. Holdout AUC was 0.848 (95% CI, 0.763-0.933), sensitivity at 70% specificity was 0.733, Brier score was 0.146, and the calibration intercept and slope were 0.988 and 2.796, respectively. For TB versus ORC, retained features were L5 consolidation, R5 consolidation, L3 consolidation, L3 B-lines with irregular pleura, and L3 B-lines with smooth pleura. Holdout AUC was 0.809 (95% CI, 0.709-0.909), sensitivity at 70% specificity was 0.690, Brier score was 0.174, and the calibration intercept and slope were -0.112 and 2.276, respectively (Figure 3, e-Tables 9-11, e-Figure 5).

**Figure 3.**
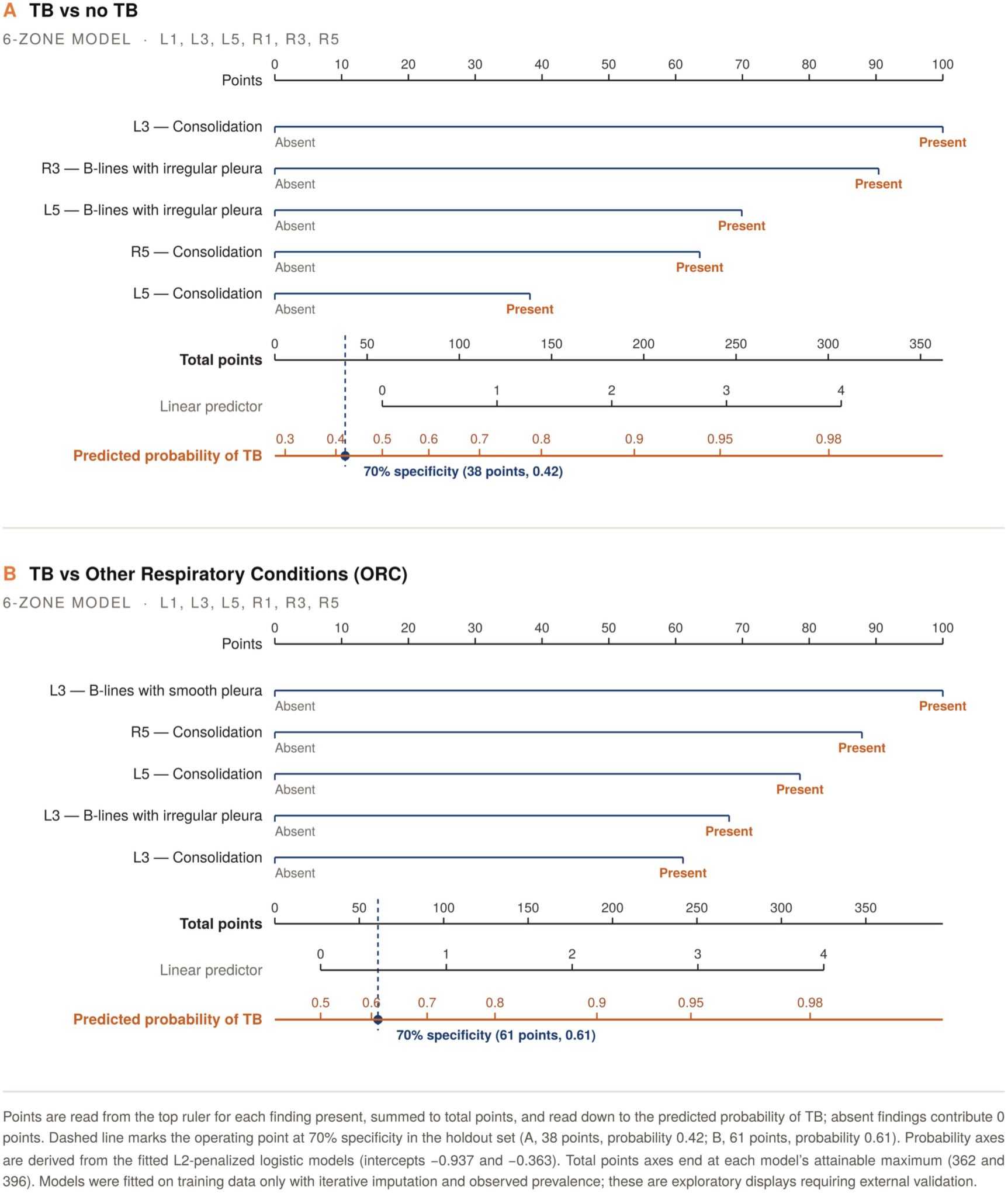
Six-zone nomograms for TB classification: L2-penalized logistic-regression models display five RF-selected POCUS features for (A) TB versus all TB-negative participants and (B) TB versus other respiratory conditions. Points assigned to present findings are summed to estimate TB probability; dashed lines indicate the thresholds corresponding to 70% specificity. These exploratory models require external validation.

## DISCUSSION

In this prospective study, a simplified 6-zone lung examination (L1, L3, L5, R1, R3, R5), derived retrospectively from the training data, performed similarly to comprehensive 12-zone and 18-zone protocols in distinguishing TB from all TB-negative participants. The comprehensive protocols performed slightly better in distinguishing TB from other respiratory conditions, although the differences were not statistically significant.

The six zones cover bilateral anterior-superior, lateral-superior, and posterior-superior lung regions where pulmonary TB commonly predominates.^20,21^ The 2-, 4-, and 6-zone solutions progressively added superior regions, supporting an anatomically coherent signal. Apical oxygen pressure, lymphatic clearance, and perfusion-to-ventilation patterns may contribute to the upper-zone predominance of post-primary TB.^20^ Other lung ultrasound studies have also associated apical and superior-zone consolidations with pulmonary TB.^14–17^ In contrast to the six-zone 12-view sweep technique, which standardizes broad apex-to-diaphragm lung acquisition for bedside interpretation and trainability,^22^ the 6-zone exam is specifically optimized for triage of microbiologically-confirmed TB.

This abbreviated 6-zone exam may reduce scanning time compared with a 12-zone scan. Its portability, lower infrastructure requirements, and absence of ionizing radiation may make it particularly useful where chest radiography is unavailable or limited for other reasons, such as costs.^8,10,18^ If validated, this may be a more desirable imaging approach for pregnant women and children and allows repeated imaging to assess for response to treatment.^8^ The six zones align closely with scanning regions in established frameworks such as the BLUE protocol^12^, which uses standardized lung ultrasound points to differentiate causes of acute respiratory failure, and E-FAST which incorporates thoracic ultrasound for the detection of pneumothorax and intrathoracic fluid^23^, which may facilitate integration into existing lung-ultrasound training and workflows.

The 6-zone strategy may not suit every clinical use case. Distinguishing TB from ORC was more difficult than distinguishing TB from TB-negative participants overall, reflecting shared sonographic findings such as consolidations and effusions.^14,16^ Comprehensive examinations may remain relevant for symptomatic patients without superior-zone abnormalities, particularly in health facilities where their greater acquisition burden may be more feasible.

Across analyses, the top zone-feature predictors were consistently pathologic findings: shred sign, consolidation, and B-lines paired with pleural irregularity. These findings reflect disruption of the normal pleural interface and parenchymal heterogeneity typical of TB granulomatous inflammation.^10,13,14,21^ Their visual distinctiveness may support rapid recognition and the future development of automated or semi-automated AI-assisted interpretation. Importantly, normal-lung signs such as the curtain sign—which reflects preserved aerated lung at the costophrenic recess^24^—were not used as risk predictors in the final feature set, because their inclusion in earlier exploratory analyses yielded risk associations inconsistent with their physiologic interpretation as markers of normal lung. Restricting the model to pathologic findings improved interpretability and aligned model-selected predictors with established sonographic markers of pulmonary TB.^10,14^

Comparison with chest radiography confirmed the expected limitation of transthoracic ultrasound. Cavity or abscess was not recorded on POCUS in any participant with radiographic cavitation.^13^ Those who were POCUS positive were identified through consolidation or subpleural nodules, nearly all in participants with peripheral infiltrates, consistent with previous reports.^10,13^ Similarly, almost all POCUS-positive participants with central infiltration had accompanying peripheral disease, and POCUS positivity was more than twice as frequent when peripheral infiltration was present. Most POCUS findings occurred in a radiographically involved lung field, although spatial correspondence does not establish that both modalities identified the same lesion. More than a quarter of participants with TB whom POCUS did not identify had a normal chest radiograph, and the remainder had less extensive radiographic disease, suggesting that a proportion of POCUS-negative TB reflect limited disease.

This study has several limitations. First, the study was conducted in a single high-burden TB country, and findings may not generalize to populations with different disease epidemiology or comorbidity profiles. Meanwhile, the model selection of apical/superior lung zones where pulmonary TB classically predominates suggests biologic relevance of the 6-zone exam across TB populations. Second, despite structured training and competency assessments, ultrasound remains operator-dependent, and inter-operator variability may have influenced feature detection. Consistent with this, eight participants with radiographic cavitation were POCUS negative, including seven with peripheral infiltration on chest radiography. ORC diagnoses were assigned clinically, which may limit conclusions about specific respiratory conditions. The analysis focused on distinguishing TB from non-TB, for which the specific ORC diagnosis was not required. Additionally, the abbreviated protocol was derived retrospectively from comprehensive examinations. Therefore, its effects on scanning time, operator burden, image quality, and diagnostic performance require prospective evaluation. Furthermore, holdout samples were modest, especially for TB versus ORC and HIV-positive or smear-negative subgroups. Sparse predictors required exclusions and penalization, so nomograms and calibration estimates remain exploratory. Both RF and nomogram models require external validation before clinical use.

Altogether, these results support a tiered approach: a fixed 6-zone scan for high-throughput case finding where chest radiography is unavailable, with comprehensive lung examination when differentiation from other respiratory conditions is the main clinical question. Prospective implementation studies should assess scanning time, training burden, cost, throughput, and confirmatory referrals. Further studies should compare ultrasound acquisition methods and feature extraction models, including fully automated deep learning approaches. These comparisons would help assess generalizability and identify opportunities to improve diagnostic performance and reproducibility.

## Ethical Approval

The study received ethical approval from the Rutgers University Institutional Review Board (eIRB #Pro2022001456) and the Mulago Hospital Research and Ethics Committee (MHREC #2349) in Uganda. All participants provided informed consent before enrollment.

## Acknowledgements

Funding was provided by the National Institute of Allergy and Infectious Diseases (NIAID), part of the U.S. National Institutes of Health (NIH). Research reported in this publication was supported by the National Cancer Institute under award numbers - R01CA249992 - 01A1, R01CA257612 - 01A1, R01CA264017 - 01, R01CA268287 - 01A1, U01CA113913 - 16A1, U01CA269181 - 01, U24CA274494 - 01; U54CA302465 - 01; R01CA268207-01; the National Heart, Lung and Blood Institute under award numbers - R01HL158071 - 01A1; the National Institute of Allergy and Infectious Diseases under award number - R01AI175555 - 01A1 and R01HL151277 - 01A1, the National Institute of Biomedical Imaging and Bioengineering under award number 75N92022D00015; the National Library of Medicine under award number - R01LM013864 - 01A1; the National Institute on Aging under award number - R01AG089759; the National Institute of Diabetes and Digestive and Kidney Diseases under award number - R01DK118431; the Office of the Director, National Institutes of Health under award number 1OT2OD038065-01; the Kidney Mapping and Atlas Project (KMAP) under award number - U01DK133090 - 01; the United States Department of Veterans Affairs VA Merit Review award - IBX004121; the VA Biomedical Laboratory Research and Development Service under award numbers - IK6BX006185; the VA Research and Development Office through the Lung Precision Oncology Program - LPOP-L0021; and sponsored research agreements from Astrazeneca, Bristol Myers Squibb, the Prevent Cancer Foundation, Innovation in Cancer Informatics, the Institute for Technology in Health Care, the Breast Cancer Research Foundation and the Scott R. Mackenzie Foundation. The content is solely the responsibility of the authors and does not necessarily represent the official views of the National Institutes of Health, the U.S. Department of Veterans Affairs, or the United States Government.

## Data Availability Statement

The data supporting the findings of this study are available from the corresponding author upon reasonable request, subject to applicable institutional and ethical approvals.

## Abbreviations

POCUS: point-of-care ultrasound
TB: tuberculosis
FASH: Focused Assessment with Sonography for HIV-associated TB
CXR: chest X-ray
IC: index case
HHC: household contact
MGIT: Mycobacteria Growth Indicator Tube
LJ: Löwenstein–Jensen culture
MTB/RIF: *Mycobacterium tuberculosis*/rifampicin resistance
NRC: no respiratory condition
ORC: other respiratory condition
AUC: area under the (receiver operating characteristic) curve
ROC: receiver operating characteristic
WHO: World Health Organization
LUS: lung ultrasound
RF: Random Forest
FDR: false discovery rate. LUS zones — Right lung: R1 anterior superior R2 anterior inferior R3 lateral superior R4 lateral inferior R5 posterior superior R6 posterior inferior. Left lung: L1 anterior superior L2 anterior inferior L3 lateral superior L4 lateral inferior L5 posterior superior L6 posterior inferior.

