## Supplementary Materials for "Streamlined point-of-care ultrasound examination for pulmonary tuberculosis"

#### Supplemental Material

This document contains supplemental methods, TB status definitions, the POCUS feature inventory, full holdout performance across protocols, the complete reduced-zone candidate search, exploratory subgroup analyses, nomogram coefficients and stability counts, and supplemental figures.

### Supplementary Methods

#### Study Population and Reference Standard

This prospective observational sub-study was conducted within the Uganda TBRU/Halting TB study (Rutgers eIRB #Pro2020003026 and #Pro2022001456). From July 2023 through March 2025, individuals aged 13 years or older were enrolled from:(1) pulmonary TB index cases (ICs), (2) household contacts (HHCs), or (3) symptomatic outpatient clinic participants who tested negative for TB and diagnosed with another respiratory condition (ORC). All participants were recruited as outpatients through KCC outpatient clinics. ICs met parent-study enrollment criteria based on symptoms consistent with pulmonary TB and microbiologic confirmation by Xpert MTB/RIF Ultra (Ultra) or a positive mycobacterial culture by either mycobacterial growth indicator tube (MGIT) or Löwenstein–Jensen (LJ). HHCs shared a dwelling or sleeping space with an IC and jointly accessed food or other essentials for at least 7 days during the 3 months preceding the IC's diagnosis. Individuals who had received more than 15 days of TB treatment during the preceding 30 days were excluded. Severely ill participants and those likely to be lost to follow-up were excluded. Final TB status for the present analysis was determined from Xpert MTB/RIF Ultra (Ultra) and mycobacterial culture results (mycobacterial growth indicator tube [MGIT] and Löwenstein–Jensen [LJ]) according to prespecified criteria. Ultra trace results were not counted as positive unless culture-confirmed (e-Table 1). Sputum-smear status was used to characterize microbiologic burden. All participants provided informed consent or, for minors, assent with parental/guardian permission.

#### POCUS Acquisition and Quality Assurance

All participants underwent a standardized 18-position point-of-care ultrasound (POCUS) examination comprising a 12-zone lung ultrasound protocol and the 6-position Focused Assessment with Sonography for HIV/TB (FASH) protocol. The lung examination sampled anterior, lateral, and posterior regions of both hemithoraces (R1-R6 and L1-L6) (Figure 1). Examinations were performed in a single session with a Philips Lumify phased-array transducer connected to an Android tablet. A 6-second cine loop was recorded at each position and uploaded to the study database.

POCUS examinations were performed by professionally qualified sonographers who received training from expert clinicians (a radiologist and a pulmonologist with expertise in ultrasonography) through successive rounds of instruction and feedback. The sonographers completed a virtual training and competency assessments in image acquisition and initial interpretation and were blinded to TB status. At each position, the sonographer recorded prespecified sonographic findings and an interpretation.

Acquisition time for each examination component was also recorded. A pulmonologist and a radiologist with ultrasound expertise independently reviewed stored cine loops while blinded to TB status. Disagreements were resolved by consensus with the sonographer. The recorded POCUS findings and their treatment in the primary Random Forest (RF) analysis are provided in e-Table 2.

#### Chest Radiography

Chest-x-rays were obtained through the parent study for ICs and HHCs. The separately enrolled ORC group did not undergo protocol-mandated chest radiography. Frontal and lateral views were interpreted using a predefined study case report form with three sections: (1) an overall interpretation (normal, findings attributable to TB, findings unlikely to represent TB, or indeterminate), (2) specific findings including central and peripheral infiltration, cavitation, pleural effusion, hilar or mediastinal lymphadenopathy, and miliary pattern, and (3) lung fields involved right or left upper and lower lung fields, defined as radiographic regions rather than anatomic lobes. A pulmonologist interpreted the images blinded to individual participant TB status and POCUS findings, but aware of the total number and proportion of pulmonary TB cases. Thus, blinding was partial.

#### Processed POCUS Feature Set

The primary lung feature matrix contained eight candidate pathologic feature families per zone: subpleural nodules, consolidation, cavity or abscess, B-lines with irregular pleura, B-lines with smooth pleura, echogenic pleural fluid, anechoic pleural fluid, and shred sign. Most were direct binary REDCap-derived fields. B-lines with irregular pleura and B-lines with smooth pleura were the only engineered composites and required both the B-lines checkbox and the corresponding pleural-line morphology checkbox to be present. No participants were coded positive for cavity or abscess on POCUS, so this variable was excluded from feature testing and model selection.

Curtain sign, A-lines, and lung sliding were recorded as normal-aeration signs but were not used as pathologic risk features. Air bronchograms, absent lung sliding, lymphadenopathy, lung point, normal lung, and standalone B-lines or pleural morphology were not independent model inputs. FASH positions were represented by their prespecified cardiac and abdominal findings.

#### Fixed Split and Reduced-Zone Selection

Random Forest (RF) classifiers were implemented in Python using scikit-learn, with the participant as the unit of analysis and final microbiologic TB status as the outcome. The two primary classification tasks were TB vs all TB-negative participants (N = 318, training n = 238, holdout n = 80, including 30 participants with TB), and TB vs ORC (N = 220,

training n = 165, holdout n = 55, including 29 participants with TB). TB vs participants with no respiratory condition (NRC) was prespecified as a supplemental analysis (N=216, training n=162, holdout n=54, including 29 participants with TB). Analyses used the complete-case, REDCap-derived processed feature matrix without imputation.
Each endpoint used one fixed stratified 75/25 training/holdout split. The same IDs were used for every protocol compared within that endpoint, with no overlap between training and holdout participants:

| Endpoint | Total N | Training N (TB+) | Holdout N (TB+) |
|---|---|---|---|
| TB vs No TB | 318 | 238 (88) | 80 (30) |
| TB vs ORC | 220 | 165 (89) | 55 (29) |
| TB vs NRC | 216 | 162 (89) | 54 (29) |

All six symmetric 2-zone, 15 symmetric 4-zone, and 20 symmetric 6-zone lung combinations were evaluated. Within each training fold, feature selection and SMOTE were performed using the fold's training portion only. Hyperparameters were fixed. Mean AUC was estimated using five-fold stratified cross-validation repeated 10 times. The highest mean cross-validation AUC within each zone-count category for TB vs No TB determined the selected 2-, 4-, and 6-zone compositions. The selected zone compositions were then applied without alteration to TB vs ORC and TB vs NRC. The holdout sets were reserved for evaluation and paired DeLong comparisons.

The selected 6-zone protocol comprised L1, L3, L5, R1, R3, and R5, corresponding to the bilateral anterior-superior, lateral-superior, and posterior-superior lung regions. RF models used 100 trees, class-weighted learning, and a random seed of 42.

#### Holdout Metrics and Subgroup Analyses

AUC confidence intervals used DeLong's method. Sensitivity at 70% specificity was interpolated from the holdout ROC curve, and 95% confidence intervals used 1,000 bootstrap samples. Precision and recall were calculated at the threshold maximizing Youden's index within the holdout predictions.

Zone-level enrichment of POCUS findings was assessed using two-sided Fisher exact tests, with Benjamini-Hochberg false discovery rate adjustment across 91 nonconstant processed LUS and FASH features.

Exploratory subgroup analyses were performed by restricting the saved holdout predictions according to HIV and sputum-smear status, without model refitting or subgroup-specific feature selection. Smear-stratified analyses retained the comparator group appropriate to the classification task and included either smear-positive or smear-negative TB cases. AUC and sensitivity at 70% specificity were summarized within each stratum. Multiple hypothesis adjustments were performed with Benjamini-Hochberg false discovery rate. These findings were considered exploratory because the HIV-positive and smear-negative strata each included only five or six TB-positive participants in the holdout sample.

#### Nomogram Modeling

Moderate nomograms were constructed separately from the RF diagnostic models using RF-selected features from the selected 6-zone protocol (L1, L3, L5, R1, R3, R5). Iterative imputation was fit on the training set only and applied to the holdout set. Nomogram fitting used the observed training prevalence without SMOTE. Complete-separation predictors were excluded, and retained features were required to be present in at least three TB-positive and three TB-negative training participants. L2-penalized logistic regression was used because sparse and quasi-separated features remained. Consequently, coefficient tables are descriptive penalized estimates. Conventional standard errors, *P* values, and odds-ratio confidence intervals are not reported.

Nomogram points were derived from the penalized regression coefficients rather than RF feature importance, and thus do not necessarily correspond to the highest-ranked RF features. Holdout performance was evaluated using AUC, sensitivity at 70% and 60% specificity, Brier score, calibration intercept, and calibration slope.

---

#### Clinical and Radiographic Statistical Analyses

Analyses were performed using Stata 18.0 (StataCorp). Continuous variables are reported as median (IQR), median (range), or mean (SD), as indicated, and compared using the Wilcoxon rank-sum test. Categorical variables are reported as number (percentage) and compared using the chi-square test, or the Fisher exact test when expected cell counts were less than 5. For variables with more than two categories, a single overall *P* value was reported. Comparisons are shown for TB versus NRC and TB versus ORC without adjustment for multiple testing. All tests were two-sided and $P < 0.05$ was considered significant. In a secondary analysis restricted to participants with protocol chest radiography, we used the same tests to compare radiographic findings in participants with pulmonary TB according to whether POCUS identified a TB-suggestive finding — subpleural nodules or masses, consolidation, cavity or abscess, miliary pattern, or pleural effusion — in any scanned zone. Anatomical correspondence was defined as such a finding in a superior zone when an upper radiographic field was involved, or an inferior zone when a lower field was involved, on the same side.

#### Notes on Excluded and Exploratory Analyses

- **Curtain sign:** Curtain sign was excluded from the primary pathologic feature set because it represents normal aerated lung at the costophrenic recess. Its earlier use as a risk feature produced clinically discordant directions.
- **Eight-zone protocols:** Eight-zone combinations appeared in earlier exploratory analyses but were not part of the canonical final candidate search. The final search comprised all symmetric 2-, 4-, and 6-zone combinations plus 12-zone and 18-position anchors.
- **FASH variables:** The final 18-position feature-selection procedure retained FASH response variables, but the 18-position AUC did not differ from the 12-zone lung AUC for either primary endpoint.
- **Expanded and clinical nomograms:** Additional all-feature, top-five, count-based, and clinical composite nomograms remain exploratory and are not the moderate nomograms displayed in the main manuscript.

### e-Tables

##### e-Table 1. Definitions of TB status according to Xpert MTB/RIF Ultra and mycobacterial culture results

*Cited in main text: Material and Methods → Study Population and Reference Standard.*

| Xpert MTB/RIF Ultra | Mycobacterial culture (MGIT and Löwenstein–Jensen) | TB status |
|---|---|---|
| Negative | One negative/contaminated/missing/unknown, or both negative | Negative |
| Negative or trace | Either positive for MTB | Positive |
| Negative | Both contaminated/missing/unknown | Negative |
| Positive or trace | Either positive for MTB | Positive |
| Positive | Contaminated/missing/unknown, or negative | Positive |
| Trace | Contaminated/missing/unknown, or negative | Indeterminate |
| Missing/error/no result | Both negative for MTB | Negative |
| Missing/error/no result | One missing/contaminated and one negative, or both contaminated/missing/unknown | Indeterminate |
| Missing/error/no result | Either positive for MTB | Positive |

Final TB status combined the Xpert MTB/RIF Ultra result with MGIT and Löwenstein–Jensen culture results. The terms *one*, *both*, and *either* refer to the two cultures considered jointly.

[a] "Positive" for Ultra refers to semiquantitative values other than "trace."

MGIT = Mycobacteria Growth Indicator Tube; MTB = *Mycobacterium tuberculosis*; TB = tuberculosis.

---

##### e-Table 2. POCUS signs and findings recorded and their role in the primary RF analysis

*Cited in main text: Material and Methods → POCUS Acquisition and Image Interpretation.*

| Sign or finding | Protocol | Interpretation | Role in primary RF analysis |
|---|---|---|---|
| A-lines | Lung | Normal aeration sign | Excluded |
| Curtain sign | Lung | Normal aeration at costophrenic recess | Excluded |
| Lung sliding | Lung | Normal pleural movement | Excluded |
| Subpleural nodules or masses | Lung | Pathologic finding | Direct binary input |
| Consolidation | Lung | Pathologic finding | Direct binary input |
| Cavity or abscess | Lung | Pathologic finding | Candidate direct input; constant in final matrix |
| Shred sign | Lung | Pathologic finding | Direct binary input |
| B-lines with irregular pleura | Lung | B-lines plus abnormal pleural morphology | Composite binary input |
| B-lines with smooth pleura | Lung | B-lines plus smooth pleural morphology | Composite binary input |
| Echogenic pleural fluid | Lung | Pathologic pleural finding | Direct binary input |
| Anechoic pleural fluid | Lung | Pathologic pleural finding | Direct binary input |

| Sign or finding | Protocol | Interpretation | Role in primary RF analysis |
|---|---|---|---|
| Standalone B-lines or pleural morphology | Lung | Artifact/morphology component | Used only to construct composites |
| Air bronchograms, absent lung sliding, lung point, standalone lymphadenopathy | Lung | Recorded pathologic signs | Not independent inputs |
| Collapse, possible miliary pattern, pleural thickening, spine sign, pneumothorax, pulmonary edema | Lung | Recorded interpretation/findings | Not independent inputs |
| Pericardial or pleural effusion, abdominal lymph nodes, hepatic lesions, splenic lesions, ascites, other/no abnormal finding | FASH | Cardiac and abdominal findings | One-hot candidate inputs in 18-position model |

FASH = Focused Assessment with Sonography for HIV-associated TB; POCUS = point-of-care ultrasound; RF = Random Forest.

---

**e-Table 3. TB versus NRC diagnostic performance**

*Cited in main text: Results → Diagnostic Performance of Reduced and Comprehensive POCUS Protocols.*

| Protocol | Zones/positions | AUC (95% CI) | Sensitivity at 70% specificity (95% CI) | Precision | Recall |
|---|---|---|---|---|---|
| 2-zone | L3, R3 | 0.677 (0.564-0.789) | 0.561 (0.397-0.711) | 0.929 | 0.448 |
| 4-zone | L3, L5, R3, R5 | 0.819 (0.716-0.922) | 0.748 (0.600-0.883) | 0.944 | 0.586 |
| 6-zone | L1, L3, L5, R1, R3, R5 | 0.797 (0.692-0.902) | 0.753 (0.608-0.881) | 0.870 | 0.690 |
| 12-zone lung | R1-R6, L1-L6 | 0.823 (0.721-0.926) | 0.759 (0.609-0.898) | 0.909 | 0.690 |
| 18-position lung+FASH | 12 lung zones + 6 FASH positions | 0.870 (0.780-0.961) | 0.852 (0.710-0.963) | 0.885 | 0.793 |

The shared TB-vs-NRC holdout set contained 54 participants, including 29 TB-positive participants. Precision and recall were calculated at the endpoint-specific Youden threshold; sensitivity at 70% specificity is a separate prespecified operating point. FASH = Focused Assessment with Sonography for HIV-associated TB; NRC = no respiratory condition.

---

**e-Table 4. Primary-endpoint holdout performance**

*Cited in main text: Results → Diagnostic Performance of Reduced and Comprehensive POCUS Protocols.*

| Endpoint | Protocol | N (TB+) | AUC (95% CI) | Sensitivity at 70% specificity (95% CI) | Precision | Recall |
|---|---|---|---|---|---|---|
| TB vs No TB | 2-zone | 80 (30) | 0.699 (0.600-0.798) | 0.594 (0.447-0.749) | 0.778 | 0.467 |

| Endpoint | Protocol | N (TB+) | AUC (95% CI) | Sensitivity at 70% specificity (95% CI) | Precision | Recall |
|---|---|---|---|---|---|---|
| TB vs No TB | 4-zone | 80 (30) | 0.771 (0.661-0.880) | 0.788 (0.652-0.906) | 0.786 | 0.733 |
| TB vs No TB | 6-zone | 80 (30) | 0.825 (0.731-0.918) | 0.814 (0.678-0.927) | 0.793 | 0.767 |
| TB vs No TB | 12-zone lung | 80 (30) | 0.810 (0.704-0.916) | 0.818 (0.689-0.926) | 0.821 | 0.767 |
| TB vs No TB | 18-position lung+FASH | 80 (30) | 0.811 (0.708-0.914) | 0.793 (0.634-0.914) | 0.778 | 0.700 |
| TB vs ORC | 2-zone | 55 (29) | 0.766 (0.660-0.872) | 0.700 (0.532-0.839) | 0.857 | 0.621 |
| TB vs ORC | 4-zone | 55 (29) | 0.812 (0.709-0.914) | 0.782 (0.621-0.903) | 0.875 | 0.724 |
| TB vs ORC | 6-zone | 55 (29) | 0.806 (0.704-0.908) | 0.754 (0.592-0.896) | 0.870 | 0.690 |
| TB vs ORC | 12-zone lung | 55 (29) | 0.849 (0.764-0.934) | 0.786 (0.655-0.903) | 0.952 | 0.690 |
| TB vs ORC | 18-position lung+FASH | 55 (29) | 0.849 (0.758-0.941) | 0.779 (0.621-0.905) | 0.952 | 0.690 |

Precision and recall were calculated at the endpoint-specific Youden threshold; sensitivity at 70% specificity is a separate prespecified operating point. ORC = other respiratory condition.

---

**e-Table 5. Selected paired DeLong AUC comparisons reported in the manuscript**

*Cited in main text: Results → Diagnostic Performance of Reduced and Comprehensive POCUS Protocols.*

| Endpoint | Model 1 (AUC) | Model 2 (AUC) | DeLong *P* value |
|---|---|---|---|
| TB vs No TB | 2-zone (0.699) | 6-zone (0.825) | **.0042** |
| TB vs No TB | 4-zone (0.771) | 6-zone (0.825) | .2172 |
| TB vs No TB | 6-zone (0.825) | 12-zone lung (0.810) | .7667 |
| TB vs No TB | 6-zone (0.825) | 18-position lung+FASH (0.811) | .7852 |
| TB vs No TB | 12-zone lung (0.810) | 18-position lung+FASH (0.811) | .9439 |
| TB vs ORC | 4-zone (0.812) | 6-zone (0.806) | .7449 |
| TB vs ORC | 6-zone (0.806) | 12-zone lung (0.849) | .4556 |
| TB vs ORC | 6-zone (0.806) | 18-position lung+FASH (0.849) | .4862 |
| TB vs ORC | 12-zone lung (0.849) | 18-position lung+FASH (0.849) | .9748 |

Comparisons are paired on the shared endpoint-specific holdout participants. The table is limited to comparisons explicitly reported in the manuscript; the complete pairwise matrix remains in the reproducibility package.

---

**e-Table 6. Candidate reduced-zone compositions evaluated for protocol selection**

*Cited in main text: Results → Diagnostic Performance of Reduced and Comprehensive POCUS Protocols.*

Selection was performed for TB vs No TB using the highest mean repeated-cross-validation AUC within each zone-count category. Holdout AUCs are shown for transparency but did not determine the selected compositions. Cross-validation intervals are the 2.5th and 97.5th percentiles across the 50 validation fits.

### Two-zone candidates

| Zones | Training CV AUC (95% interval) | Holdout AUC (95% CI) | Selected |
|---|---|---|---|
| L3, R3 | 0.665 (0.556-0.789) | 0.699 (0.600-0.798) | Yes |
| L1, R1 | 0.622 (0.496-0.747) | 0.780 (0.682-0.878) | No |
| L2, R2 | 0.615 (0.480-0.711) | 0.698 (0.604-0.792) | No |
| L5, R5 | 0.600 (0.493-0.676) | 0.786 (0.691-0.881) | No |
| L6, R6 | 0.535 (0.464-0.617) | 0.546 (0.467-0.625) | No |
| L4, R4 | 0.520 (0.459-0.567) | 0.469 (0.407-0.530) | No |

### Four-zone candidates

| Zones | Training CV AUC (95% interval) | Holdout AUC (95% CI) | Selected |
|---|---|---|---|
| L3, L5, R3, R5 | 0.701 (0.583-0.814) | 0.771 (0.661-0.880) | Yes |
| L2, L3, R2, R3 | 0.696 (0.596-0.793) | 0.764 (0.665-0.863) | No |
| L1, L3, R1, R3 | 0.695 (0.555-0.830) | 0.801 (0.703-0.900) | No |
| L1, L2, R1, R2 | 0.681 (0.559-0.824) | 0.788 (0.674-0.902) | No |
| L3, L4, R3, R4 | 0.676 (0.588-0.801) | 0.707 (0.610-0.804) | No |
| L3, L6, R3, R6 | 0.663 (0.570-0.762) | 0.673 (0.555-0.790) | No |
| L2, L5, R2, R5 | 0.659 (0.521-0.787) | 0.744 (0.628-0.860) | No |
| L1, L5, R1, R5 | 0.658 (0.518-0.785) | 0.861 (0.776-0.947) | No |
| L1, L4, R1, R4 | 0.635 (0.485-0.747) | 0.771 (0.665-0.878) | No |
| L2, L4, R2, R4 | 0.631 (0.496-0.735) | 0.661 (0.552-0.770) | No |
| L2, L6, R2, R6 | 0.622 (0.463-0.739) | 0.706 (0.603-0.809) | No |
| L1, L6, R1, R6 | 0.621 (0.476-0.738) | 0.819 (0.723-0.914) | No |
| L4, L5, R4, R5 | 0.610 (0.517-0.705) | 0.793 (0.701-0.886) | No |
| L5, L6, R5, R6 | 0.581 (0.472-0.662) | 0.753 (0.641-0.864) | No |
| L4, L6, R4, R6 | 0.539 (0.441-0.647) | 0.515 (0.427-0.602) | No |

### Six-zone candidates

| Zones | Training CV AUC (95% interval) | Holdout AUC (95% CI) | Selected |
|---|---|---|---|
| L1, L3, L5, R1, R3, R5 | 0.718 (0.570-0.827) | 0.825 (0.731-0.918) | Yes |
| L2, L3, L5, R2, R3, R5 | 0.718 (0.623-0.834) | 0.771 (0.670-0.871) | No |
| L3, L4, L5, R3, R4, R5 | 0.710 (0.560-0.816) | 0.771 (0.663-0.879) | No |
| L1, L3, L4, R1, R3, R4 | 0.706 (0.572-0.838) | 0.813 (0.721-0.906) | No |
| L1, L2, L5, R1, R2, R5 | 0.705 (0.559-0.837) | 0.826 (0.726-0.926) | No |
| L2, L3, L4, R2, R3, R4 | 0.705 (0.611-0.779) | 0.769 (0.672-0.866) | No |
| L3, L5, L6, R3, R5, R6 | 0.704 (0.604-0.807) | 0.742 (0.629-0.855) | No |
| L1, L2, L3, R1, R2, R3 | 0.701 (0.573-0.829) | 0.816 (0.713-0.919) | No |

| Zones | Training CV AUC (95% interval) | Holdout AUC (95% CI) | Selected |
|---|---|---|---|
| L2, L3, L6, R2, R3, R6 | 0.694 (0.576-0.775) | 0.716 (0.597-0.836) | No |
| L1, L3, L6, R1, R3, R6 | 0.692 (0.575-0.813) | 0.791 (0.681-0.901) | No |
| L1, L2, L4, R1, R2, R4 | 0.689 (0.574-0.821) | 0.716 (0.589-0.843) | No |
| L2, L4, L5, R2, R4, R5 | 0.679 (0.556-0.824) | 0.745 (0.631-0.859) | No |
| L1, L2, L6, R1, R2, R6 | 0.673 (0.539-0.788) | 0.747 (0.620-0.874) | No |
| L1, L4, L5, R1, R4, R5 | 0.671 (0.509-0.793) | 0.859 (0.775-0.943) | No |
| L3, L4, L6, R3, R4, R6 | 0.670 (0.570-0.781) | 0.684 (0.570-0.798) | No |
| L2, L5, L6, R2, R5, R6 | 0.653 (0.465-0.760) | 0.734 (0.612-0.856) | No |
| L1, L5, L6, R1, R5, R6 | 0.652 (0.512-0.758) | 0.816 (0.707-0.924) | No |
| L1, L4, L6, R1, R4, R6 | 0.633 (0.479-0.744) | 0.783 (0.674-0.892) | No |
| L2, L4, L6, R2, R4, R6 | 0.627 (0.492-0.725) | 0.683 (0.570-0.796) | No |
| L4, L5, L6, R4, R5, R6 | 0.593 (0.505-0.690) | 0.704 (0.583-0.825) | No |

The selected 6-zone (1/3/5 zones) candidate had a marginally higher mean training cross-validation AUC than 2/3/5 zones (0.718232 versus 0.718182); both round to 0.718. Additional training-only cross-validation analyses performed subsequently also modestly favored 1/3/5 over this runner-up: sensitivity at 70% specificity was 0.651 versus 0.648, balanced accuracy at a fixed threshold of 0.50 was 0.704 versus 0.697, and balanced accuracy at the Youden threshold was 0.731 versus 0.723. The small differences do not establish meaningful superiority of the selected composition and underscore the need for external validation.

---

**e-Table 7. Exploratory subgroup analyses by HIV status**

*Cited in main text: Results → Exploratory Subgroup Analyses.*

| Endpoint | HIV stratum | Protocol | N | TB+ | TB– | AUC (95% CI) | Sensitivity at 70% specificity (95% CI) |
|---|---|---|---|---|---|---|---|
| TB vs No TB | HIV-positive | 6-zone | 13 | 5 | 8 | 0.925 (0.766-1.000) | 1.000 (0.840-1.000) |
| TB vs No TB | HIV-positive | 12-zone lung | 13 | 5 | 8 | 0.725 (0.364-1.000) | 0.847 (0.460-1.000) |
| TB vs No TB | HIV-positive | 18-position lung+FASH | 13 | 5 | 8 | 0.725 (0.364-1.000) | 0.816 (0.400-1.000) |
| TB vs No TB | HIV-negative | 6-zone | 67 | 25 | 42 | 0.802 (0.695-0.909) | 0.778 (0.627-0.904) |
| TB vs No TB | HIV-negative | 12-zone lung | 67 | 25 | 42 | 0.828 (0.721-0.936) | 0.812 (0.663-0.930) |
| TB vs No TB | HIV-negative | 18-position lung+FASH | 67 | 25 | 42 | 0.828 (0.721-0.934) | 0.789 (0.622-0.929) |
| TB vs ORC | HIV-positive | 6-zone | 15 | 5 | 10 | 0.760 (0.467-1.000) | 0.650 (0.200-1.000) |
| TB vs ORC | HIV-positive | 12-zone lung | 15 | 5 | 10 | 0.880 (0.679-1.000) | 0.850 (0.526-1.000) |

| Endpoint | HIV stratum | Protocol | N | TB+ | TB− | AUC (95% CI) | Sensitivity at 70% specificity (95% CI) |
|---|---|---|---|---|---|---|---|
| TB vs ORC | HIV-positive | 18-position lung+FASH | 15 | 5 | 10 | 0.900 (0.702-1.000) | 0.850 (0.526-1.000) |
| TB vs ORC | HIV-negative | 6-zone | 40 | 24 | 16 | 0.819 (0.704-0.934) | 0.782 (0.635-0.916) |
| TB vs ORC | HIV-negative | 12-zone lung | 40 | 24 | 16 | 0.854 (0.765-0.943) | 0.781 (0.653-0.905) |
| TB vs ORC | HIV-negative | 18-position lung+FASH | 40 | 24 | 16 | 0.839 (0.730-0.947) | 0.756 (0.577-0.910) |

Models were not refit. Estimates use the saved fixed-holdout predictions and should be interpreted cautiously because the HIV-positive strata contained five TB-positive participants.

---

**e-Table 8. Exploratory subgroup analyses by sputum smear status**

*Cited in main text: Results → Exploratory Subgroup Analyses.*

| Endpoint | TB stratum | Protocol | N | TB+ | TB− | AUC (95% CI) | Sensitivity at 70% specificity (95% CI) |
|---|---|---|---|---|---|---|---|
| TB vs No TB | Smear-positive TB | 6-zone | 74 | 24 | 50 | 0.816 (0.713-0.919) | 0.801 (0.644-0.934) |
| TB vs No TB | Smear-positive TB | 12-zone lung | 74 | 24 | 50 | 0.809 (0.699-0.919) | 0.803 (0.642-0.933) |
| TB vs No TB | Smear-positive TB | 18-position lung+FASH | 74 | 24 | 50 | 0.806 (0.696-0.916) | 0.778 (0.588-0.922) |
| TB vs No TB | Smear-negative TB | 6-zone | 56 | 6 | 50 | 0.860 (0.685-1.000) | 0.867 (0.586-1.000) |
| TB vs No TB | Smear-negative TB | 12-zone lung | 56 | 6 | 50 | 0.813 (0.537-1.000) | 0.876 (0.607-1.000) |
| TB vs No TB | Smear-negative TB | 18-position lung+FASH | 56 | 6 | 50 | 0.833 (0.573-1.000) | 0.856 (0.542-1.000) |
| TB vs ORC | Smear-positive TB | 6-zone | 50 | 24 | 26 | 0.835 (0.730-0.940) | 0.802 (0.635-0.933) |
| TB vs ORC | Smear-positive TB | 12-zone lung | 50 | 24 | 26 | 0.840 (0.746-0.933) | 0.770 (0.626-0.892) |
| TB vs ORC | Smear-positive TB | 18-position lung+FASH | 50 | 24 | 26 | 0.840 (0.738-0.942) | 0.767 (0.583-0.900) |
| TB vs ORC | Smear-negative TB | 6-zone | 31 | 5 | 26 | 0.665 (0.395-0.936) | 0.525 (0.154-1.000) |
| TB vs ORC | Smear-negative TB | 12-zone lung | 31 | 5 | 26 | 0.892 (0.713-1.000) | 0.862 (0.557-1.000) |
| TB vs ORC | Smear-negative TB | 18-position lung+FASH | 31 | 5 | 26 | 0.896 (0.692-1.000) | 0.840 (0.477-1.000) |

All endpoint-appropriate TB-negative comparators were retained, and TB-positive participants were restricted by smear status. Models were not refit. Smear-negative strata contained six TB-positive participants for TB vs No TB and five for TB vs ORC.

**e-Table 9. Moderate nomogram performance and retained predictors**

*Cited in main text: Results → Clinically-interpretable Nomograms.*

| Endpoint | Protocol | Retained predictors | AUC (95% CI) | Sn at 70% Sp | Brier score | Calibration intercept | Calibration slope |
|---|---|---|---|---|---|---|---|
| TB vs No TB | 6-zone | L3 consolidation; L5 consolidation; L5 B-lines with irregular pleura; R3 B-lines with irregular pleura; R5 consolidation | 0.848 (0.763-0.933) | 0.733 | 0.146 | 0.988 | 2.796 |
| TB vs ORC | 6-zone | L5 consolidation; R5 consolidation; L3 consolidation; L3 B-lines with irregular pleura; L3 B-lines with smooth pleura | 0.809 (0.709-0.909) | 0.690 | 0.174 | -0.112 | 2.276 |

Both models used L1, L3, L5, R1, R3, and R5 as the source protocol. Calibration was evaluated on the fixed holdout set; no bootstrap optimism correction was performed.

**e-Table 10. Moderate nomogram coefficients, odds ratios, and points**

*Cited in main text: Results → Clinically-interpretable Nomograms.*

| Endpoint | Predictor | Penalized coefficient | Odds ratio | Points when present |
|---|---|---|---|---|
| TB vs No TB | L3 consolidation | 1.608 | 4.994 | 100.0 |
| TB vs No TB | L5 consolidation | 0.614 | 1.847 | 38.2 |
| TB vs No TB | L5 B-lines with irregular pleura | 1.125 | 3.079 | 69.9 |
| TB vs No TB | R3 B-lines with irregular pleura | 1.454 | 4.281 | 90.4 |
| TB vs No TB | R5 consolidation | 1.022 | 2.780 | 63.6 |
| TB vs ORC | L5 consolidation | 1.055 | 2.872 | 78.6 |
| TB vs ORC | R5 consolidation | 1.180 | 3.256 | 87.9 |
| TB vs ORC | L3 consolidation | 0.820 | 2.271 | 61.1 |
| TB vs ORC | L3 B-lines with irregular pleura | 0.912 | 2.490 | 68.0 |
| TB vs ORC | L3 B-lines with smooth pleura | 1.342 | 3.827 | 100.0 |

The reference state is feature absent and receives 0 points. Estimates come from L2-penalized logistic regression; conventional inferential confidence intervals and *P* values are not applicable to these displayed penalized coefficients.

**e-Table 11. Feature-present 2×2 stability counts for retained nomogram predictors**

*Cited in main text: Results → Clinically-interpretable Nomograms.*

| Endpoint | Predictor | TB+ present | TB– present | TB+ absent | TB– absent |
|---|---|---|---|---|---|
| TB vs No TB | L3 consolidation | 18 | 3 | 70 | 147 |

| Endpoint | Predictor | TB+ present | TB– present | TB+ absent | TB– absent |
|---|---|---|---|---|---|
| TB vs No TB | L5 consolidation | 11 | 4 | 77 | 146 |
| TB vs No TB | L5 B-lines with irregular pleura | 10 | 3 | 78 | 147 |
| TB vs No TB | R3 B-lines with irregular pleura | 13 | 3 | 75 | 147 |
| TB vs No TB | R5 consolidation | 13 | 5 | 75 | 145 |
| TB vs ORC | L5 consolidation | 16 | 3 | 73 | 73 |
| TB vs ORC | R5 consolidation | 17 | 3 | 72 | 73 |
| TB vs ORC | L3 consolidation | 16 | 3 | 73 | 73 |
| TB vs ORC | L3 B-lines with irregular pleura | 15 | 3 | 74 | 73 |
| TB vs ORC | L3 B-lines with smooth pleura | 15 | 3 | 74 | 73 |

Counts are from the training sets before nomogram fitting. The moderate screening rule required at least three TB-positive and three TB-negative feature-present participants.

**e-Table 12. Chest radiograph findings according to POCUS result among 117 participants with pulmonary tuberculosis**

| Chest Radiograph Finding | POCUS Positive (n = 81) | POCUS Negative (n = 36) | P Value |
|---|---|---|---|
| ***Specific radiographic findings*** | | | |
| Central infiltration | 64 (79.0) | 18 (50.0) | .002 |
| Peripheral infiltration | 74 (91.4) | 22 (61.1) | < .001 |
| Cavitation | 30 (37.0) | 8 (22.2) | .137 |
| Pleural effusion | 9 (11.1) | 2 (5.6) | .499 |
| Hilar or mediastinal lymphadenopathy | 14 (17.3) | 7 (19.4) | .798 |
| Miliary pattern | 1 (1.2) | 0 (0.0) | 1.000 |
| Other finding | 12 (14.8) | 3 (8.3) | .389 |
| Normal radiograph | 2 (2.5) | 10 (27.8) | < .001 |
| ***Distribution of disease*** | | | |
| Any upper lung field | 73 (90.1) | 24 (66.7) | .003 |
| Any lower lung field | 64 (79.0) | 15 (41.7) | < .001 |
| Both upper and lower fields | 58 (71.6) | 13 (36.1) | < .001 |
| Lung fields involved, mean (SD) | 2.4 (1.0) | 1.6 (1.3) | < .001 |
| ***Overlapping radiographic findings*** | | | |
| Central + peripheral infiltration | 61 (75.3) | 14 (38.9) | < .001 |
| Central infiltration + cavitation | 26 (32.1) | 6 (16.7) | .116 |
| Peripheral infiltration + cavitation | 29 (35.8) | 7 (19.4) | .087 |
| Central + peripheral + cavitation | 25 (30.9) | 5 (13.9) | .067 |

| Chest Radiograph Finding | POCUS Positive (n = 81) | POCUS Negative (n = 36) | P Value |
|---|---|---|---|
| More than one radiographic finding | 69 (85.2) | 18 (50.0) | < .001 |
| Radiographic findings, mean (SD) | 2.5 (1.0) | 1.7 (1.4) | .002 |
| ***Peripheral disease*** | | | |
| Peripheral infiltration present (n = 96) | 74 (77.1) | — | < .001 |
| Peripheral infiltration absent (n = 21) | 7 (33.3) | — | |
| ***Correspondence between modalities*** | | | |
| Cavity or abscess recorded on POCUS | 0 (0.0) | 0 (0.0) | — |
| Lymphadenopathy recorded on POCUS | 0 (0.0) | 0 (0.0) | — |
| POCUS finding in a radiographically involved field | 78/81 (96.3) | — | — |
| Upper-level concordance, of those with upper field involved | 70/73 (95.9) | — | — |
| Lower-level concordance, of those with lower field involved | 45/64 (70.3) | — | — |

Data are No. (%) unless otherwise indicated. POCUS positive denotes a TB-suggestive finding — subpleural nodules or masses, consolidation, cavity or abscess, miliary pattern, or pleural effusion — recorded in any of 12 scanned zones. Comparisons used the Fisher exact test for proportions and the Wilcoxon rank-sum test for continuous variables. Lung fields and radiographic findings are presented as mean (SD). Rows are not mutually exclusive. One TB participant did not undergo CXR due to pregnancy and was excluded. For POCUS positivity by presence of peripheral disease, denominators are all 117 participants stratified by whether peripheral infiltration was present on radiography; values are the proportion within each stratum who were POCUS positive, and the P value compares the two strata.

Cavity or abscess and lymphadenopathy were not recorded on POCUS in any participant, including the 38 with radiographic cavitation and the 21 with hilar or mediastinal lymphadenopathy. Among the 30 participants with cavitation who were POCUS positive, positivity was attributable to consolidation (n = 20) or subpleural nodules (n = 12), or both (n = 2), and 29 of 30 also had peripheral infiltration on radiography. No participant had cavitation as the sole radiographic finding.

Concordance denotes a TB-suggestive POCUS finding in a zone corresponding to a radiographically involved lung field. Lung fields are radiographic zones on the frontal projection, not anatomic lobes. Participants with other respiratory conditions did not undergo protocol chest radiography and are not included. POCUS = point-of-care ultrasound.

### e-Figures

##### e-Figure 1. Participant flow diagram

eFigure 1  Participant flow

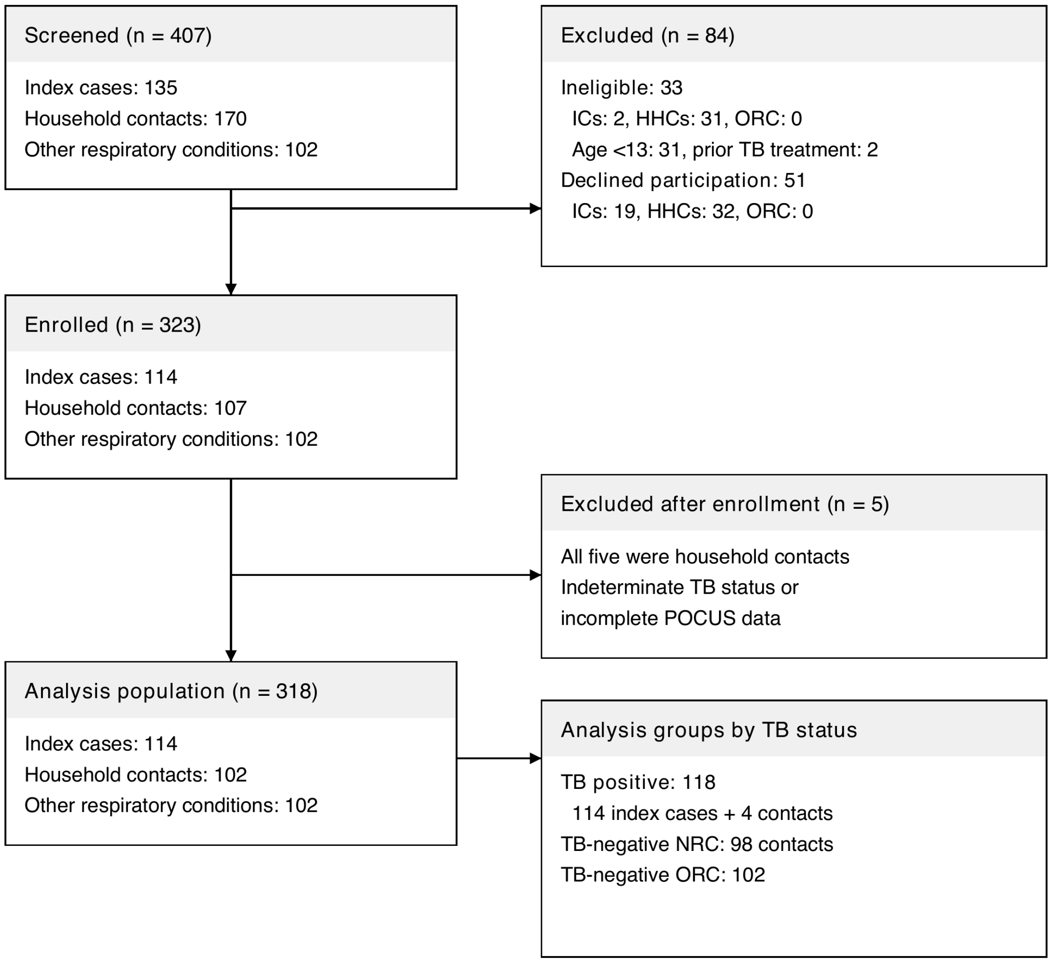


IC, index case. HHC, household contact.
TB, tuberculosis. NRC, no respiratory condition. ORC, other respiratory condition.
POCUS, point-of-care ultrasound.

Flow from 407 screened participants through 323 enrolled participants to the final analysis population of 318 participants. Excluded (n = 84): 33 ineligible (31 aged < 13 y, 2 prior anti-tuberculosis treatment per protocol) and 51 declined. The final cohort included 114 index cases, 102 household contacts, and 102 symptomatic clinic evaluees with ORC; 118 were TB-positive, 98 were TB-negative NRC, and 102 were TB-negative ORC.

**e-Figure 2. Selected 6-zone RF feature importance for TB versus all TB-negative participants**

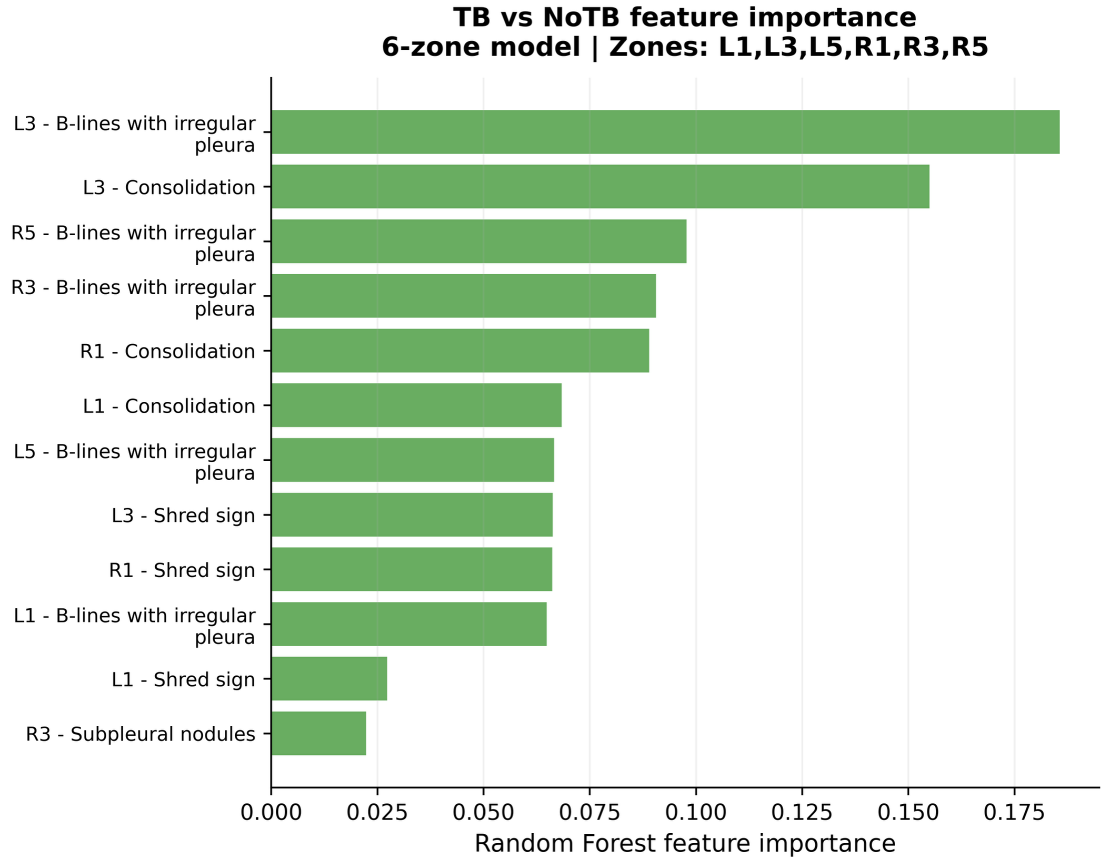


Random Forest feature importance within the selected L1/L3/L5/R1/R3/R5 protocol for TB versus all TB-negative participants.

**e-Figure 3. Selected 6-zone RF feature importance for TB versus NRC**

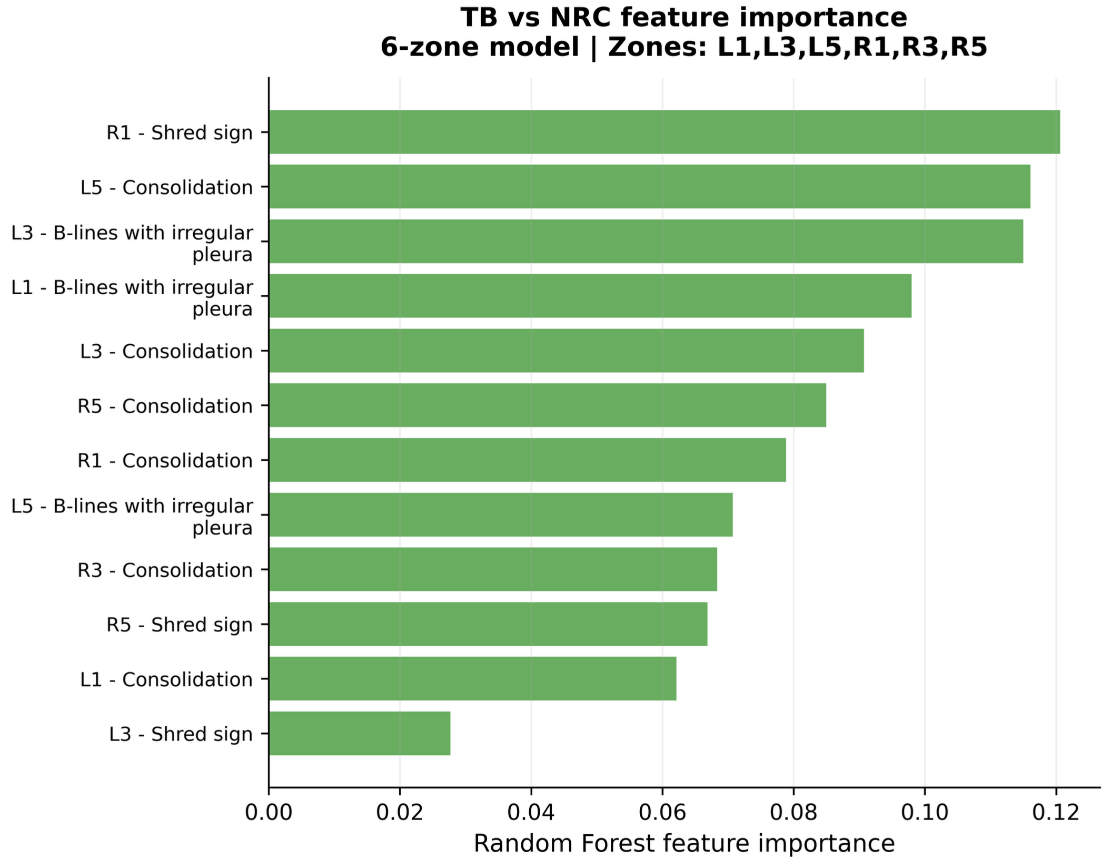


Random Forest feature importance within the selected L1/L3/L5/R1/R3/R5 protocol for TB versus participants without a respiratory condition.

**e-Figure 4. Selected 6-zone RF feature importance for TB versus ORC**

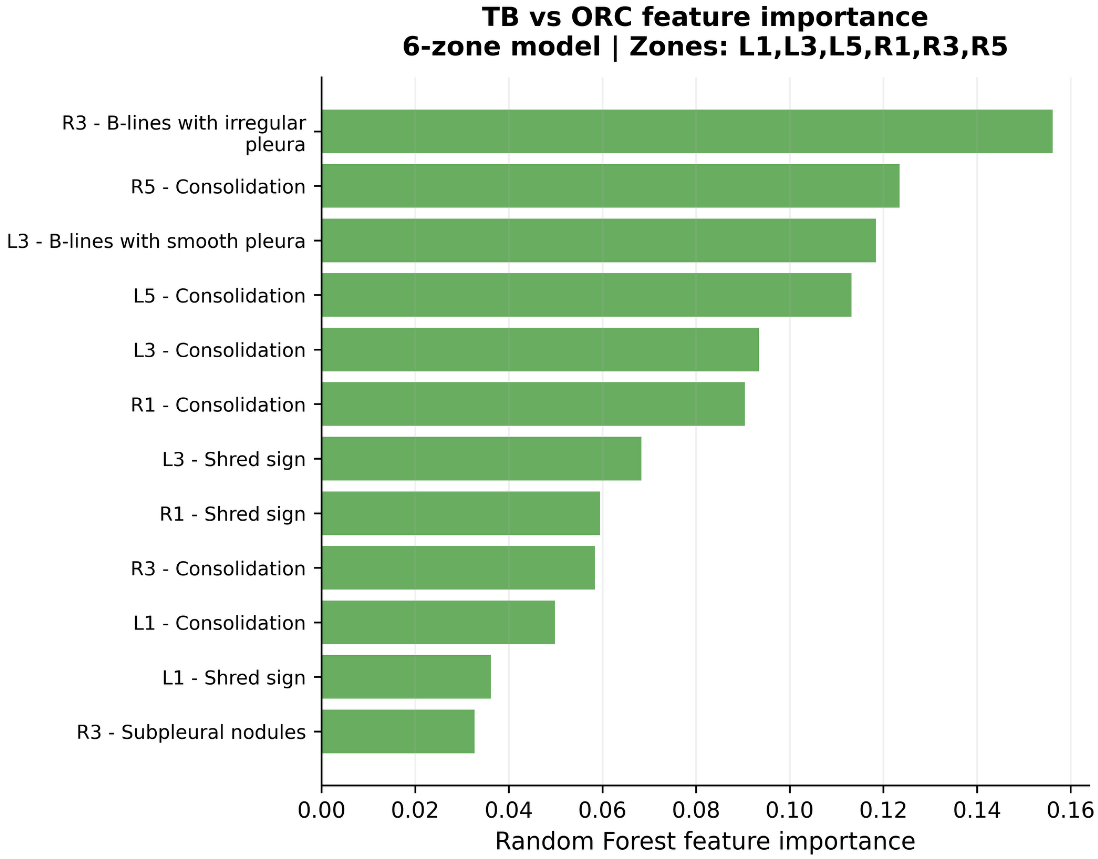


Random Forest feature importance within the selected L1/L3/L5/R1/R3/R5 protocol for TB versus participants with other respiratory conditions.

**e-Figure 5. Moderate nomogram holdout calibration**

**A. TB versus all TB-negative participants**

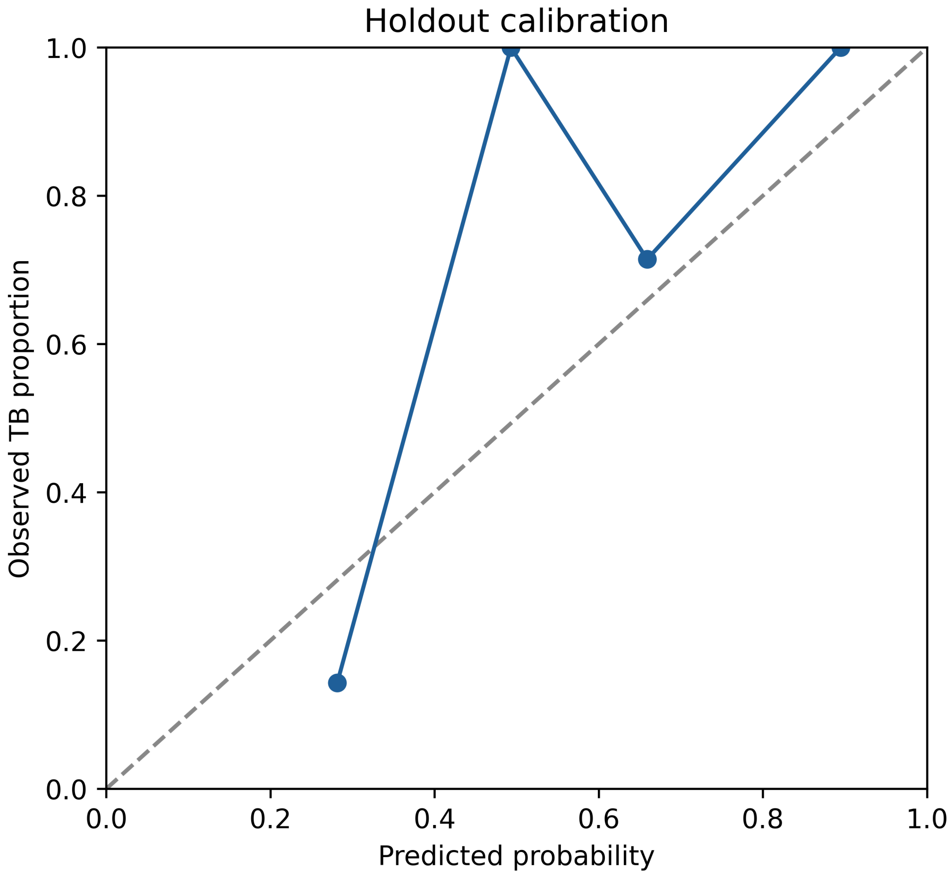

**B. TB versus ORC**

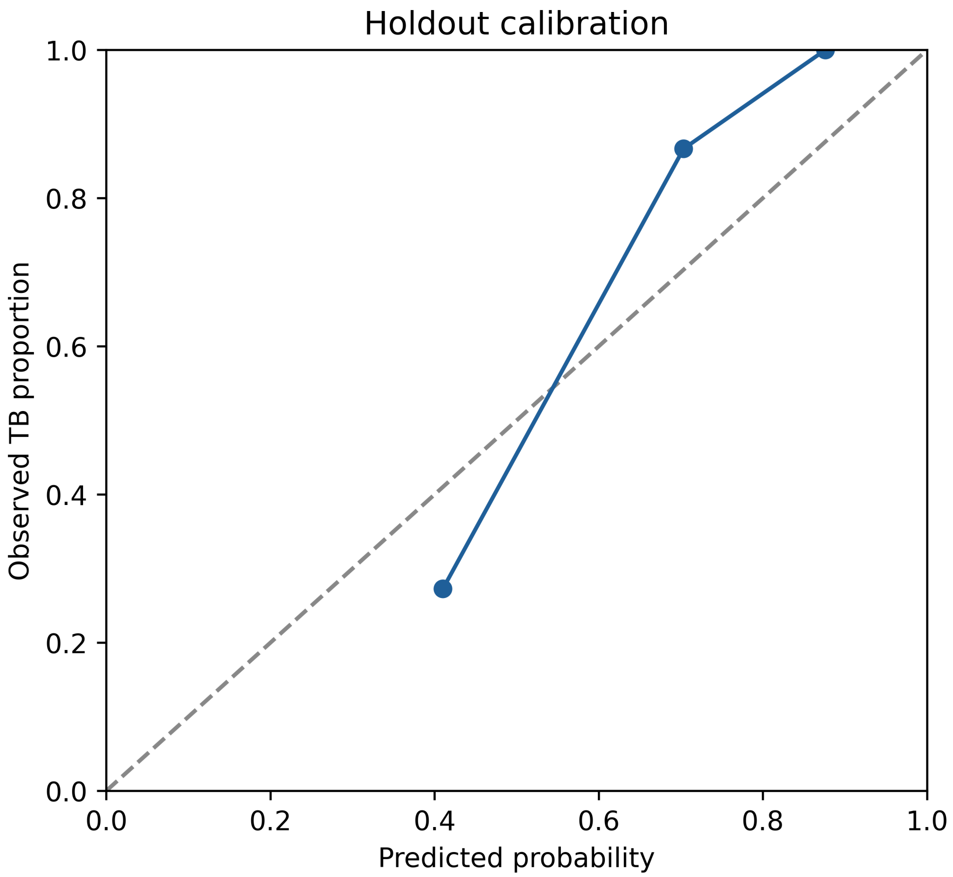


Observed TB proportions are plotted against mean predicted probability in quantile groups. The diagonal denotes ideal calibration. Calibration estimates should be interpreted cautiously because of the small fixed holdout sets.
